# Cortical differences across psychiatric disorders and associated common and rare genetic variants

**DOI:** 10.1101/2025.04.16.25325971

**Authors:** Kuldeep Kumar, Zhijie Liao, Jakub Kopal, Clara Moreau, Christopher R. K. Ching, Claudia Modenato, Will Snyder, Sayeh Kazem, Charles-Olivier Martin, Anne-Marie Bélanger, Valérie K. Fontaine, Khadije Jizi, Rune Boen, Guillaume Huguet, Zohra Saci, Leila Kushan, Ana I. Silva, 16p11.2 European Consortium, Simons Searchlight Consortium, Marianne B.M. van den Bree, David E.J. Linden, Michael J. Owen, Jeremy Hall, Sarah Lippé, Guillaume Dumas, Bogdan Draganski, Laura Almasy, Sophia I. Thomopoulos, Neda Jahanshad, Ida E. Sønderby, Ole A. Andreassen, David C. Glahn, Armin Raznahan, Carrie E. Bearden, Tomas Paus, Paul M. Thompson, Sébastien Jacquemont

## Abstract

Genetic studies have identified common and rare variants increasing the risk for neurodevelopmental and psychiatric disorders (NPDs). These risk variants have also been shown to influence the structure of the cerebral cortex. However, it is unknown whether cortical differences associated with genetic variants are linked to the risk they confer for NPDs. To answer this question, we analyzed cortical thickness (CT) and surface area (SA) for common and rare variants associated with NPDs, in ∼33000 individuals from the general population and clinical cohorts, as well as ENIGMA summary statistics for 8 NPDs. Rare and common genetic variants increasing risk for NPDs were preferentially associated with total SA, while NPDs were preferentially associated with mean CT. Larger effects on mean CT, but not total SA, were observed in NPD medicated subgroups. At the regional level, genetic variants were preferentially associated with effects in sensorimotor areas, while NPDs showed higher effects in association areas. We show that schizophrenia- and bipolar-disorder-associated SNPs show positive and negative effect sizes on SA suggesting that their aggregated effects cancel out in additive polygenic models. Overall, CT and SA differences associated with NPDs do not relate to those observed across individual genetic variants and may be linked with critical non-genetic factors, such as medication and the lived experience of the disorder.

## Introduction

Neurodevelopmental and psychiatric disorders (NPDs) represent a significant burden on global health, characterized by complex etiologies rooted in brain development and function^1–4^. NPDs are highly heritable, with a broad spectrum of common and rare genetic variants implicated in their pathogenesis^2,5–13^. The increasing availability of large-scale magnetic resonance imaging (MRI) and genetics datasets has afforded the opportunity to discover genetic variants, both common and rare, influencing the structure of the human cerebral cortex^14–20^. Studies have shown that cortical morphology measures, such as cortical thickness (CT) and surface area (SA), are highly heritable^14,17,18^, are driven by distinct neurobiological processes^21–23^, and are influenced by largely separate genetic factors^14,17,18,24,25^. Consequently, considerable research has explored the genetic overlap between variants associated with NPDs and those influencing cortical morphology^14,17,18,26–29^, revealing small genetic correlations between NPDs and SA, and even weaker overlaps for CT.

In parallel to genetic investigations, large scale case-control neuroimaging studies have consistently identified cortical differences associated with a range of psychiatric disorders^30,31^. Cross-disorder analyses have further revealed shared regional patterns of cortical variation^32–37^, often aligning along the well-established sensorimotor-association cortical gradient^18,38–40^. Cortical differences have also been reported for NPD-associated rare genetic variants, including copy number variations (CNVs)^16,37,41–45^. Neuroimaging of these NPD-associated CNVs has demonstrated, on average, much larger effects on CT and SA^41–43,46^, than those observed in NPDs^15,42,43^. However, a critical question remains unanswered: whether the cortical differences associated with both common and rare genetic variants are related to the risk they confer for psychiatric disorders. The lack of large-scale cohorts integrating genetics and neuroimaging data across healthy controls and individuals with psychiatric disorders has hindered direct investigation of this crucial relationship. Therefore, to address this knowledge gap, we designed an analytical approach to systematically compare the neuroimaging signatures of common and rare genetic variants that increase the risk for psychiatric disorders to the neuroimaging signatures associated with NPDs.

Here we aggregated multiple datasets as well as published summary statistics from ENIGMA^31^ consortium to compare clinical diagnoses-related (8 NPDs) and gene-related (common and rare variants) case-control group differences on global and regional CT and SA. Specifically, we aimed to compare effect sizes on CT and SA for three primary categories: i) 8 NPDs^31^ (attention deficit hyperactivity disorder (ADHD)^47^; autism spectrum disorder (ASD)^48,49^; bipolar disorder (BD)^50^; clinical high-risk for psychosis (CHR-PS)^51^; conduct disorder (CD)^52^; major depressive disorder (MDD)^53^; obsessive-compulsive disorder (OCD)^54^; and schizophrenia (SCZ)^55^), considering medicated and unmedicated subgroups where available; ii) common variants^56–59^ associated with NPDs; and iii) 18 different CNV and aneuploidy rare variants associated with NPDs. Overall, we provide the first systematic evaluation of cortical differences associated with NPDs, their genetic variants (both common and rare), and place these findings in the context of twin and SNP heritability estimates^14^. These comprehensive analyses reveal a notable pattern: while psychiatric disorders preferentially affect mean CT, and association cortical regions, genetic variants impact total-SA and sensorimotor cortical regions.

## Results

### Effects of psychiatric disorders and associated genetic variants on global cortical metrics

We sought to quantify effect sizes of neurodevelopmental and psychiatric disorders (NPDs) and associated genetic variants on total cortical surface area (total SA) and mean cortical thickness (mean CT). Effect sizes (Cohen’s *d*) on total SA were an order of magnitude larger for rare genetic variants compared with NPDs (11-fold, Wilcoxon rank-sum test, FDR q<0.05, **Figure 2B**). This difference in effect sizes was less pronounced for mean CT (3-fold, Wilcoxon rank-sum test, FDR q<0.05, **Figure 2A**). Rare genetic variants were associated with larger effect sizes on total SA compared to mean CT while NPD-diagnosis showed preferential association with mean CT (paired *t*-test and ratio of Cohen’s *d*, FDR q<0.05, **Figure 2C-D**). Of note, some early onset (childhood/adolescence subgroups) conditions showed preferential effects on total SA (conduct disorder, and MDD-young, **Figure 2C**), but we did not observe a preferential effect on surface when stratifying for pediatric/young NPDs (**Supplement Figure 1**). A sensitivity analysis showed similar mean CT and total SA effect sizes for rare variants across deletions, duplications, and sex chromosome aneuploidies (**Supplement Figure 2**).

**Figure 1.**
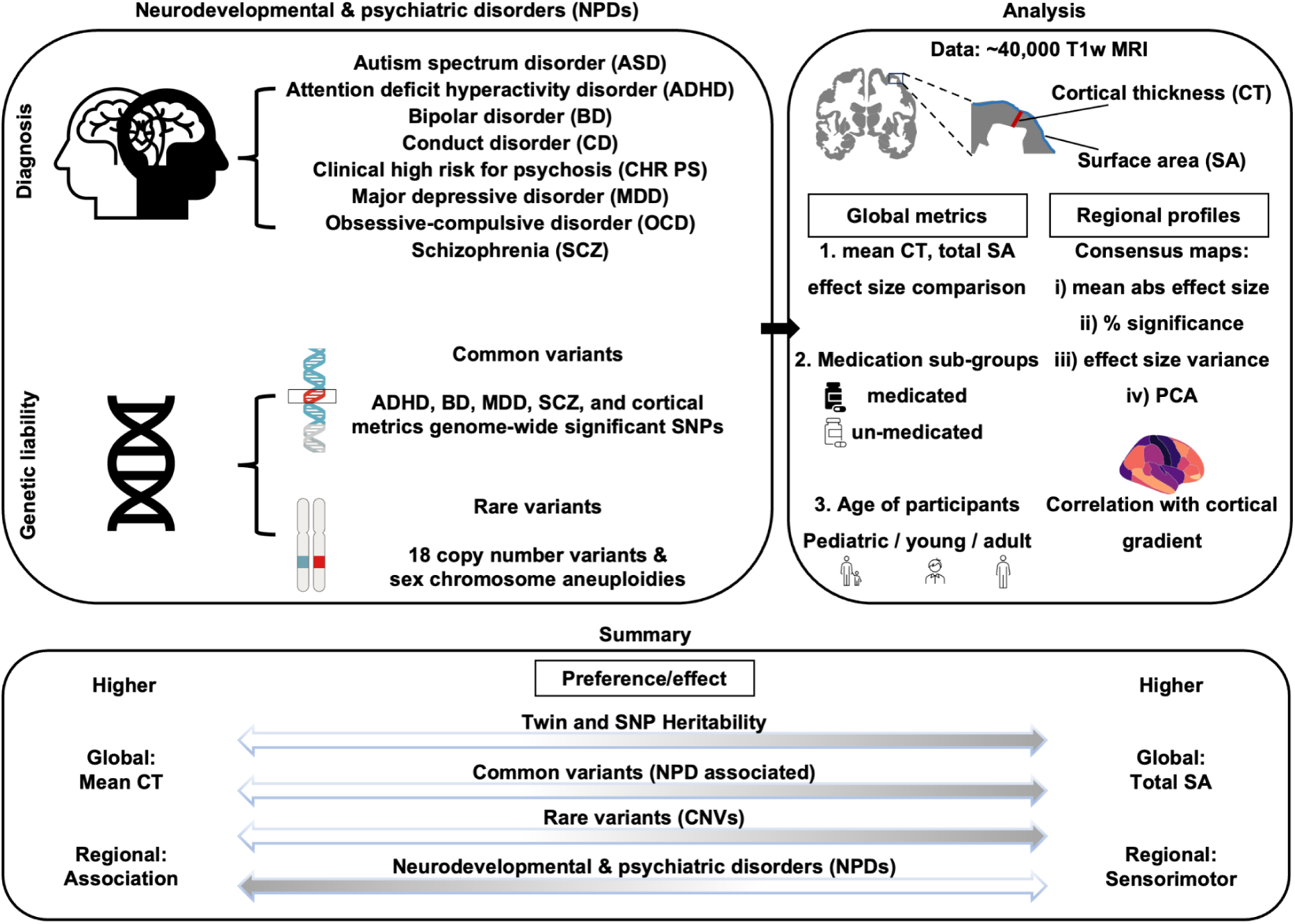
Graphical abstract. Study overview. We aimed to compare effect sizes on CT and SA for three primary categories: i) 8 NPDs^31^ (attention deficit hyperactivity disorder (ADHD)^47^; autism spectrum disorder (ASD)^48,49^; bipolar disorder (BD)^50^; clinical high-risk for psychosis (CHR-PS)^51^; conduct disorder (CD)^52^; major depressive disorder (MDD)^53^; obsessive-compulsive disorder (OCD)^54^; and schizophrenia (SCZ)^55^), considering medicated and unmedicated subgroups where available; ii) common variants^56–59^ associated with NPDs; and iii) 18 different CNV and aneuploidy rare variants associated with NPDs. Towards this, we aggregated multiple datasets as well as published summary statistics from ENIGMA^31^ consortium to compare case control effect sizes (Cohen’s d) across 8 psychiatric disorders and associated common and rare variants on global and regional CT and SA. Global and regional effect sizes were compared, and spatial patterns of variations were evaluated using four sets of consensus maps including: mean absolute effect size maps, significance maps, variance maps, and principal component analysis. Our findings across genetic variants increasing the risk for NPDs align with twin and SNP heritability estimates^14^, which are higher for surface area and sensorimotor regions, suggesting that these are general properties of the genetic architecture of the cerebral cortex. Overall, our study suggests that the neuroimaging alterations observed in NPDs are distinct from those observed across genetic variants increasing the risk for NPDs. Brain and cortex maps were generated using the *ggseg* package in R^78^. Common and rare variant illustrations are from the NIAID NIH BIOART Source (https://bioart.niaid.nih.gov/bioart/170 and https://bioart.niaid.nih.gov/bioart/204)

**Figure 2:**
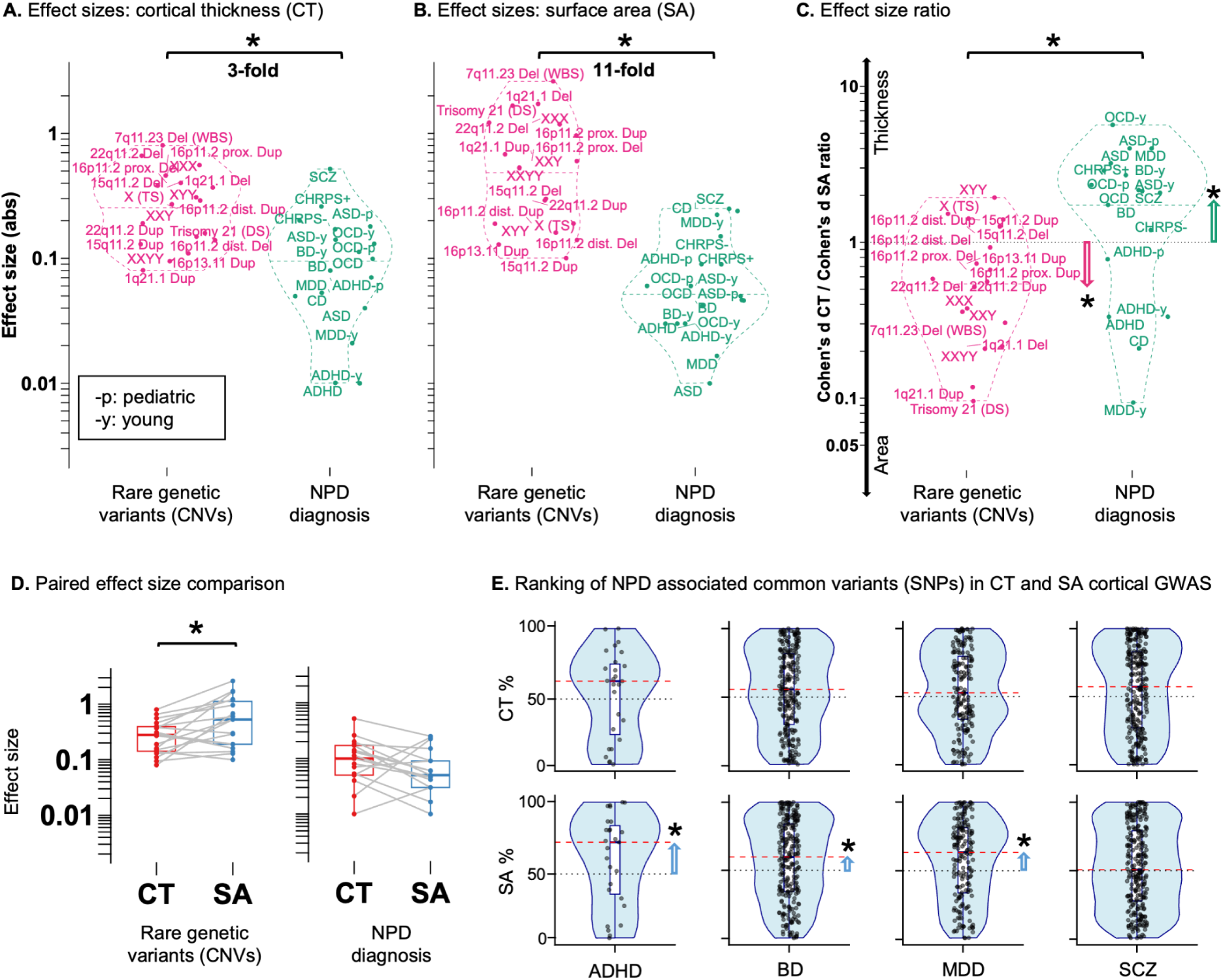
Global cortical differences across NPDs and associated genetic variants. Legend: **A-D**) Comparison between neurodevelopmental and psychiatric disorders (NPDs) associated rare genetic liability (CNVs) and diagnosis on **A**) mean cortical thickness (CT) effect sizes; **B**) total surface area (SA) effect sizes; **C**) the ratio of CT and SA effect sizes; and **D**) paired CT and SA effect sizes. Case-control differences were adjusted for age, sex, and site. *: FDR significant (q<0.05), across all pairs of comparisons. Absolute effect sizes (Y-axis) are plotted on a log10 scale. **E**) Effects on CT and SA of common variants associated with NPDs. We tested if NPD genome-wide significant SNPs were enriched in SNPs associated with SA or CT. We ranked independent NPD-associated SNPs based on their p-value association with CT and SA. Median rankings are indicated using dotted red lines. * and arrows represent significant (FDR q<0.05) median ranking compared to permutation-based null distribution. Abbreviations: Adult-Adolescence-Pediatric sample abbreviations: -y=young; -p=pediatric; Abs=absolute; ADHD=attention deficit hyperactivity disorder; ASD=autism spectrum disorder; BD=bipolar disorder; CD: conduct disorder; CHRPS: clinical high risk for psychosis; CHRPSn: CHR who did not develop a psychotic disorder; CHRPSp: CHR who later developed a psychotic disorder; CNV=copy number variant; CT=cortical thickness; Del=deletion; Dup=duplication; GWAS: genome-wide association study; MDD=major depressive disorder; NPD=neurodevelopmental and psychiatric disorders; OCD=obsessive-compulsive disorder; prox.=proximal; SA=surface area; SCZ=schizophrenia; SNP=single nucleotide polymorphism; TS=Turner syndrome; WBS=Williams-Beuren syndrome;

We then asked if preferential effects on total SA also applied to common variants associated with NPDs (ADHD^57^, BD^56^, MDD^58^, and SCZ^59^). Previous work has shown that genetic contribution to total SA was weakly correlated with NPDs. Genetic correlation between CT and NPDs has not been detected^14^. We therefore sought to identify, beyond genetic correlation, the potential genetic overlap between NPDs and cortical structure by computing a non-parametric enrichment. NPD-associated SNPs were mildly enriched (above-median ranking) in SA-associated SNPs (non-parametric p-value<0.0001; **Figure 2E**). In contrast, none of the NPD-associated SNPs showed any enrichment for association with CT. Similarly, the SA-genome-wide associated SNPs, but not CT, were also enriched in NPD associations (above-median ranking, **Supplement Figure 3**).

We performed sensitivity analyses to test if differences observed above between genetic variants and psychiatric diagnoses could be in part influenced by medication (also a proxy for severity^60^) and the age of participants (a proxy for severity or neurodevelopmental processes in earlier onset participants, as well as duration of illness in adults). Effect sizes on mean CT were 3.4-fold larger in medicated compared to unmedicated NPD subgroups (FDR q<0.05, **Figure 3A**), while total SA remained the same in both subgroups (**Figure 3B**). As a result, medicated sub-groups exhibited larger effects on mean CT compared to total SA (FDR q<0.05), while unmedicated sub-groups had similar effect sizes for both metrics **Figure 3C**). The preferential effect on mean CT observed for NPDs could not be explained by stratifying participants according to age (pediatric, young, and adult participants, **Supplement Figure 1**).

**Figure 3:**
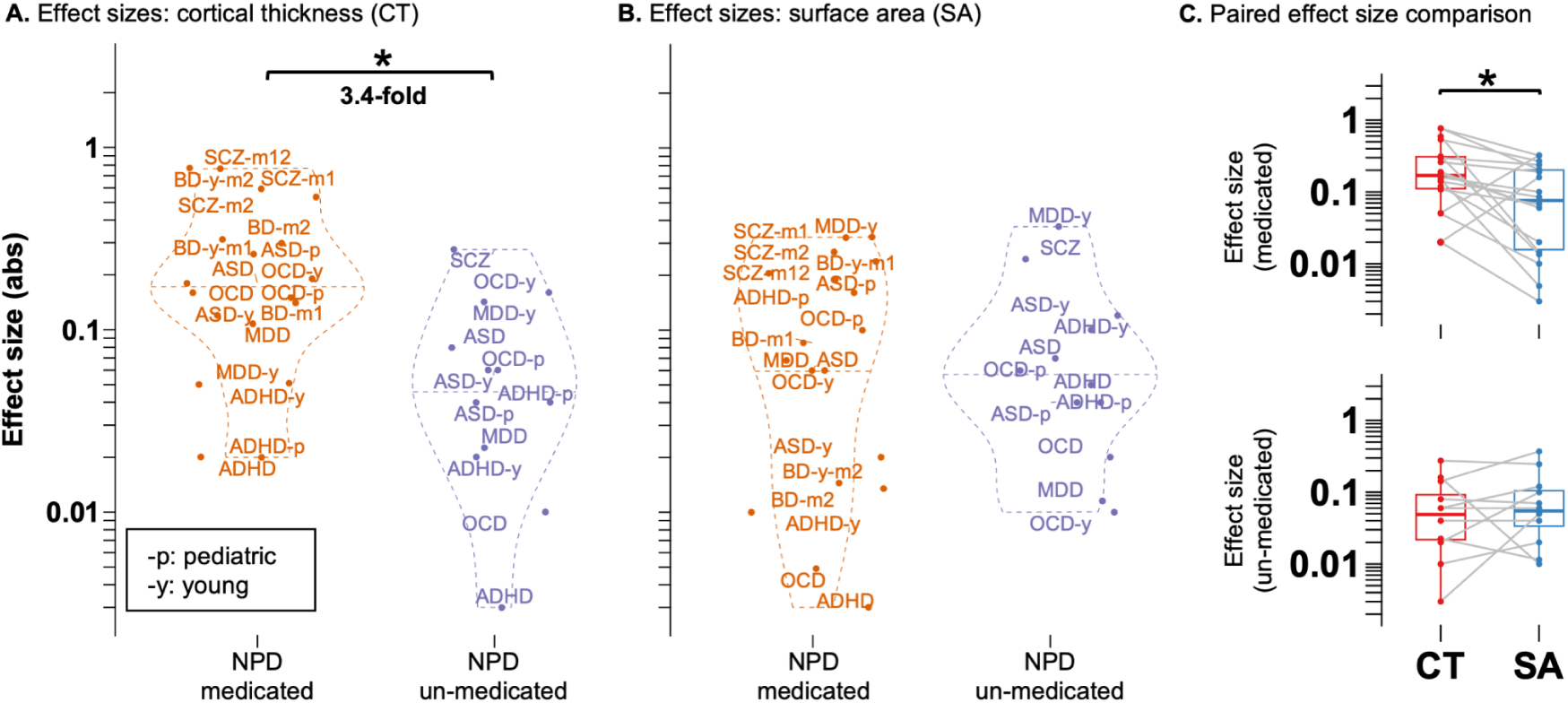
Global cortical differences across medication sub-groups. Legend: **A-C**) Comparing the effect sizes on mean cortical thickness (CT) and total surface area (SA) of neurodevelopmental and psychiatric disorders (NPDs) diagnosis sub-groups with and without medications using A-B) violin plots; and C) paired boxplots. Case-control differences were adjusted for age, sex, and site. *: FDR significant (q<0.05), across all pairs of comparisons. Absolute effect sizes (Y-axis) are plotted on a log10 scale. Abbreviations: Adult-Adolescence-Pediatric sample abbreviations: -y=young; -p=pediatric; Abs=absolute; ADHD=attention deficit hyperactivity disorder; ASD=autism spectrum disorder; BD=bipolar disorder; CD: conduct disorder; CHRPS: clinical high risk for psychosis; CHRPSn: CHR who did not develop a psychotic disorder; CHRPSp: CHR who later developed a psychotic disorder; CT=cortical thickness; Del=deletion; Dup=duplication; DZ=di-zygotic; MDD=major depressive disorder; MZ=mono-zygotic; NPD=neurodevelopmental and psychiatric disorders; OCD=obsessive-compulsive disorder; SA=surface area; SCZ=schizophrenia; Medication abbreviations: -m=medicated; -u=un-medicated; BD-m1=lithium medication; BD-m2=antiepileptics medication; SCZ-m1=1st generation; SCZ-m2=2nd generation; SCZ-m12=1st & 2nd generation;

### Opposing regional patterns of cortical differences between NPDs and genetic variants

We investigated if the preferential effects on total SA for genetic variants and mean CT for psychiatric diagnoses were uniformly distributed across the cerebral cortex or localized to specific cortical regions. To contextualize the regional profiles, we tested their similarities with the well-established cortical gradient (**Methods**), which ranks regions from primary sensory-motor cortices, to higher order association cortices^38,39^. We first examined how this gradient relates to both twin and SNP heritability estimates for regional CT and SA^14^. Heritabilities were higher in sensorimotor regions compared to association regions (negative correlations with the cortical gradient, r=-0.47 to -0.75, p-spin<0.05, **Figure 4B, Supplement Figure 4**).

**Figure 4.**
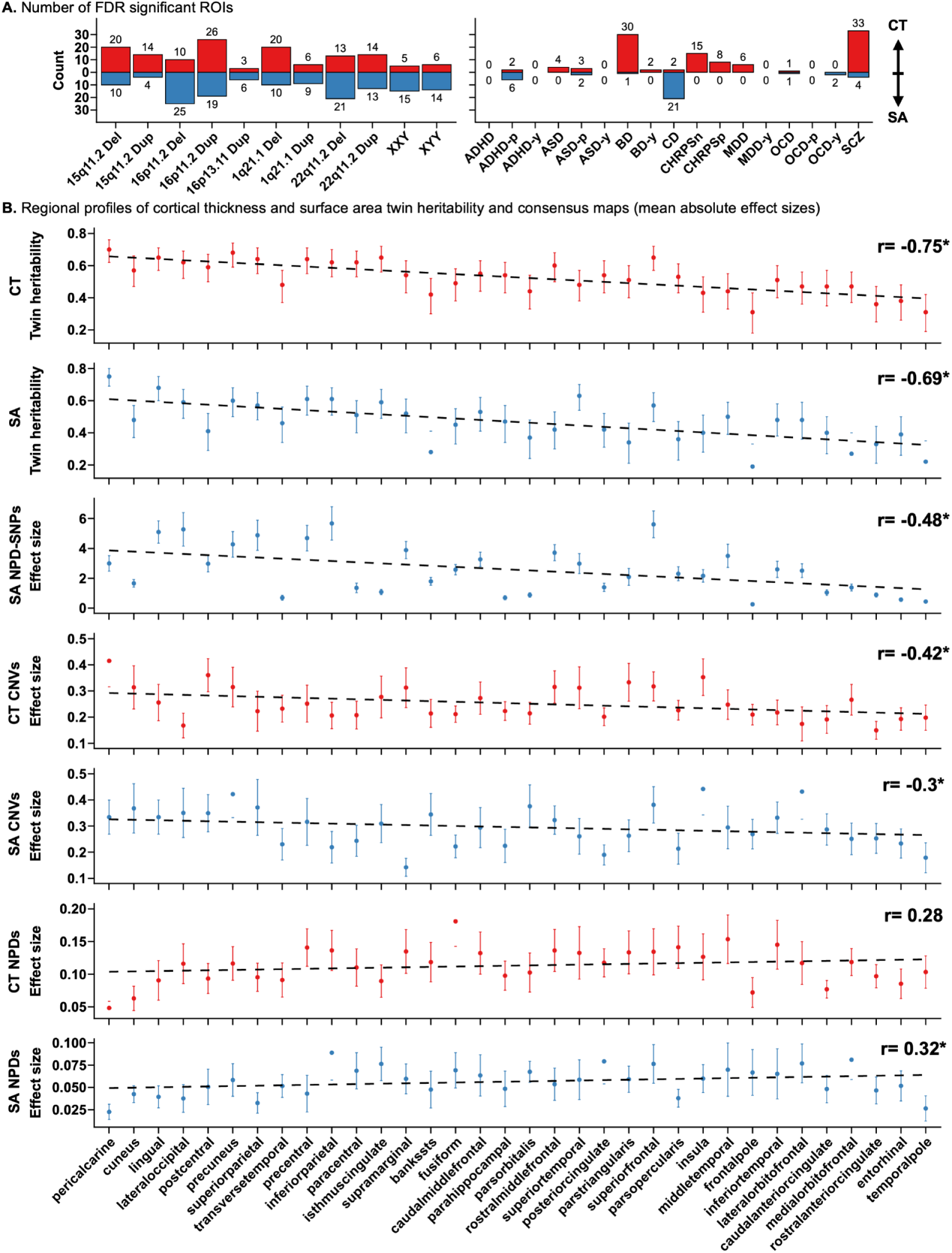
Consensus maps of regional cortical differences. Legend: **A**) Number of FDR significant cortical regions (out of 34) per CNV/NPD for cortical thickness (up, red) and surface area (down, blue). **B**) Regional profiles of twin heritability, and mean absolute effect sizes across common and rare genetic variants and NPDs for cortical thickness and surface area across 34 Desikan cortical regions. Each point represents: i) First two rows: the twin heritability and 95% CI; ii) third row: mean estimate from linear regression for NPD associated common variants (SA NPD-SNPs); and iii) bottom four rows: mean absolute effect size (Cohen’s d), with error bars showing the standard error of the mean. Y-axis: heritability estimates or effect sizes. X-axis: cortical regions ordered according to the cortical gradient from sensorimotor to association regions. Dotted line: correlation with the cortical gradient. Each panel displays Pearson correlation and *:spin-permutation significant, p-spin < 0.05. Abbreviations, Abs=absolute; ADHD=attention deficit hyperactivity disorder; ASD=autism spectrum disorder; BD=bipolar disorder; CD: conduct disorder; CHRPS: clinical high risk for psychosis; CHRPSn: CHR who did not develop a psychotic disorder; CHRPSp: CHR who later developed a psychotic disorder; CNV=copy number variant; CT=cortical thickness; Del=deletion; Dup=duplication; MDD=major depressive disorder; NPD=neurodevelopmental and psychiatric disorders; OCD=obsessive-compulsive disorder; SA=surface area; SCZ=schizophrenia; TS=Turner syndrome; Adult-Adolescence-Pediatric abbreviations: -y=adolescence/young; -p=pediatric).

We then investigated the regional cortical effect size maps of all NPDs examined above, 257 genome-wide NPD-associated SNPs^14^, and 11 CNVs (**Supplement Table 1)**. Significant associations with regional SA or CT were observed for 18 NPDs, 20 NPD-associated common variants (SNPs), and all 11 CNVs (**Figure 4A**). Cohen’s *d* maps without any FDR-significant ROI association were excluded from the regional analyses below (non-significant maps included medications subgroups, **Figure 4A, Supplement Figure 5)**.

We computed 4 set of consensus cortical maps, including 3 disregarding the directionality of effects, using following approaches: i) mean absolute effect size; ii) percentage of significance; iii) variance in effect size values; and iv) latent dimensions of cortical differences using principal component analysis across effect size profiles (**Supplement Figure 6-7**). The rare and common variant maps showed positive correlations (r=0.29 to 0.31, p-spin < 0.05, **Supplement Figure 8**), and did not show correlation with the NPD maps (**Supplement Figure 8**). NPD-associated rare and common genetic variants showed higher effect sizes in sensorimotor regions across all consensus map methods compared to association regions (**Figure 4B, 5C**, correlations with the cortical gradient: r=-0.43, p-spin=9.5e-3, for CT; r=-0.3, p-spin=2e-2, for SA). NPDs maps showed the opposite correlations with the cortical gradient highlighting larger effects in association regions (**Figure 4B, 5C**, CT: r=0.28, p-spin=9.3e-2; SA: r=0.32, p-spin=3.2e-2). Overall, NPD-associated common and rare variant effect sizes as well as twin and SNP heritability estimates for cortical measures were higher in sensorimotor regions, while the opposite was observed for NPDs (**Figure 4-5**).

**Figure 5.**
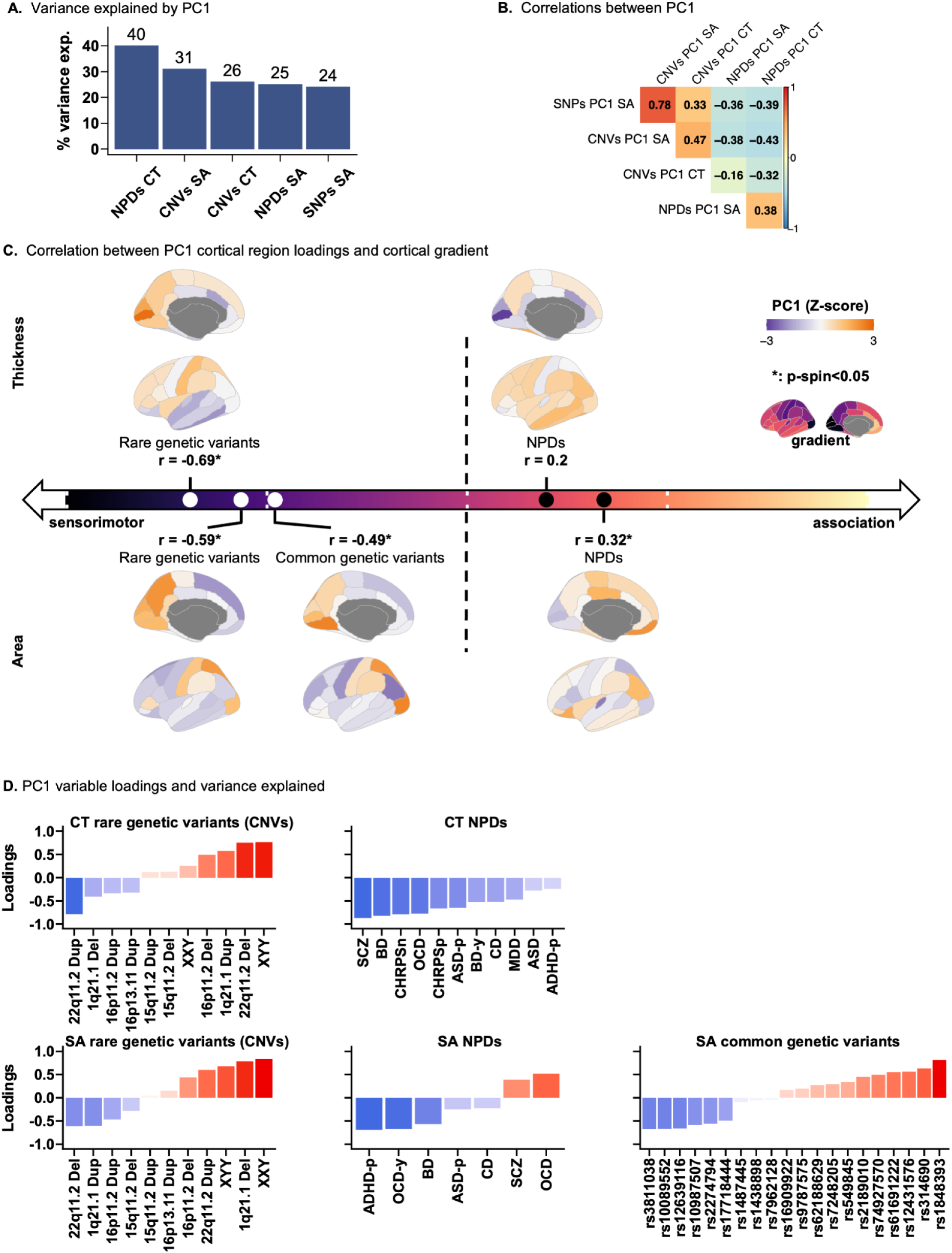
Latent dimensions of regional cortical differences. Legend: **A**) The variance explained by the first principal component (PC1) for neurodevelopmental and psychiatric disorders (NPDs), their associated common and rare genetic variants. **B**) Correlation between latent dimensions. Pairwise spatial correlations between PC1 for CT and SA across NPDs, and associated common and rare genetic variants. **C**) Correlation between the cortical gradient and the latent dimension of cortical differences. Plots are arranged along the sensorimotor-association axis^38^ based on correlation with cortical gradient (AHBA gene expression principal component). Positive and negative correlation values indicate greater similarity with association and sensorimotor cortical regions, respectively. Each plot displays Pearson correlation and *:spin-permutation significant, p-spin < 0.05. **D**) PC1 variable loadings and variance explained for NPDs and genetic variants. Abbreviations, Abs=absolute; ADHD=attention deficit hyperactivity disorder; ASD=autism spectrum disorder; BD=bipolar disorder; CD: conduct disorder; CHRPS: clinical high risk for psychosis; CHRPSn: CHR who did not develop a psychotic disorder; CHRPSp: CHR who later developed a psychotic disorder; CNV=copy number variant; CT=cortical thickness; Del=deletion; Dup=duplication; MDD=major depressive disorder; NPD=neurodevelopmental and psychiatric disorders; OCD=obsessive-compulsive disorder; PC: principal component; p-spin: spin permutation based p-value; r: Pearson correlation; SA: surface area; SCZ=schizophrenia; SNPs: single nucleotide polymorphism; TS=Turner syndrome; % variance exp.= percentage of variance explained;

### Polygenic and familial liability to NPDs

We asked if the results showing preferential associations with total SA for individual rare and common genetic variants computed above may be reconciled with the difficulty to detect associations between SA and NPDs, as well as polygenic or familial liability to NPDs. For global metrics, polygenic risk scores (PRS), PRS-BD and PRS-SCZ, were not associated with either metric in 31,000 UK Biobank participants of European ancestry (**Supplement Figure 9**). To investigate the lack of PRS signal, we used regression estimates derived from cortical structure GWAS. Half of the BD and SCZ associated SNPs had a negative beta estimate for total SA (BD: 51.05%, and SCZ: 48.26%, **Supplement Figure 9**), while the other half showed positive beta estimate, likely resulting in cancellation of the effects of additive psychiatric PRS models on cortical structure. Using summary statistics^61^ in first degree relatives, a proxy for multifactorial polygenic liability, we observed similar effects on total SA for the family member and those with a diagnosis of SCZ or BD (**Supplement Figure 10**).

To understand why genetic variants show preferential effects on sensorimotor regions for surface area, while NPDs demonstrate little or none, we examined the loadings of genetic variants on the first latent dimension. Half of the rare variants loaded positively, while the other loaded negatively on the SA latent dimension (i.e., increased and decreased surface in sensorimotor regions, **Figure 5D**). The same was true for common variants (**Figure 5D)**. As a result, we were unable to detect associations between PRS for BD and SCZ and surface area of sensorimotor regions (**Supplement Figure 9**).

## Discussion

Individual genetic variants and NPD diagnoses showed, on average, opposing effects on global cortical differences, where rare and common genetic variants associated with NPDs preferentially affected total SA while the diagnosis of NPDs was associated with mean CT. The effect of NPDs on mean CT was larger in medication sub-groups, which are proxies for disorder severity. Beyond global effects, psychiatric diagnoses preferentially affected association cortical regions (involved in higher-order functions such as language, decision-making, and social cognition) while rare and common genetic variants impacted sensorimotor cortical regions (involved in basic sensory and motor functions). We also show that the polygenic architecture of NPDs (BD and SCZ) together with the positive and negative effects sizes of genetic variants on cortical strucutre may leads to the cancellation of most of the effects of NPD polygenic scores on cortical structure.

The preferential effects on total SA, compared to mean CT, of NPD-associated rare and common variants is concordant with the higher heritability estimates for total SA compared to mean CT^14^. The observed preferential effect of NPDs on mean CT may be attributable to critical non-genetic factors including medication, environmental factors, and plasticity in the context of a psychiatric disorder. While directly modeling these effects remains challenging, our findings suggest a potential role for medication^62^, proxy for disorder severity. This also aligns with longitudinal findings from a randomized clinical trial in MDD showing medication-related changes in mean CT but not total SA^62^. Studies of familial risk, also support this notion, where individuals with either SZ or BD exhibited larger effect sizes on mean CT compared to their unaffected first degree relatives, while effects on total SA were not significant in both groups^61^. Furthermore, comparing 22q11.2 deletion carriers with and without Psychosis^45^, showed differences in mean CT but not total SA. These differential effects may be consistent with the distinct developmental trajectories of cortical SA and CT^24,25^. While the majority of cortical expansion occurs before the first two years^24,63^ and is relatively stable thereaftere^23,64^, CT undergoes protracted maturation involving changes in dendritic arborization and myelination that continue into adulthood^24,38,63–65^. As a result, further investigation of childhood onset conditions should reveal larger association with SA.

Rare and common genetic variants preferentially impact sensorimotor cortical regions, which is concordant with higher twin and SNP heritability estimates for SA and CT in those same regions^14^. In contrast, NPDs tend to affect association regions (subserving executive, socioemotional, and mentalizing functions), which is in line with previous studies^33–35^. Theses preferential effects in association areas (focal reduction in CT) have also been reported in 22q11.2 deletion carriers with a diagnosis of psychosis compared to those without^45^. These differential regional effects in individual genetic variants and those with psychiatric disorders may be consistent with studies showing that the spatiotemporal patterns of CT and SA maturation follow a sensorimotor-to-association gradient, where sensorimotor cortical regions mature before the association regions^24,38,63^.

Studies reported higher twin and SNP heritability estimates for SA compared to CT, and this study suggests the preferential effects of NPD-associated common and rare genetic variants on SA. However, detecting effects on SA in psychiatric conditions has been challenging^31^, raising a paradox given the high heritability estimates of psychiatric disorders^1,5^. Notably, some early onset (childhood/adolescence subgroups) conditions showed preferential effects on total-SA (Conduct Disorder, and MDD-young) but detecting effects on SA remains challenging for other highly heritable childhood disorders such as ASD and ADHD. Future studies of pediatric NPDs are required to shed light on these questions.

Challenges to identify effects on SA may be a consequence of polygenic architecture of NPDs. Our investigation of individual SZ- and BD-associated common variants and their aggregated effects on total and regional SA (i.e., in the sensorimotor regions) using polygenic risk scores (PRS) suggests that many of the SA effects of individual SNPs are cancelled out in a PGS model. The effects of individual variants on total and regional SA (in the sensorimotor regions) may instead point to a hidden underlying mechanism indirectly related to SA. As an example, the level of transcriptomic differences observed in autistic brains (compared to controls) follows the cortical gradient, with the larger levels of differentially expressed genes occurring in the sensorimotor compared to association regions^66^.

There are several limitations of our study. We are only investigating 8 NPDs and selected recurrent CNVs, this is largely due to the limited availability of the large CNV MRI cohorts, as well as robust associations of these CNVs with other conditions with published summary statistics^43,67^. While our analysis showed similar observations across sex chromosome aneuploidies and CNVs, we have to note that aneuploidies have specific effects during puberty, and which will require careful delineation^64,68^. Finally, the ENIGMA medication analyses were not always able to account for confounding factors like illness severity and duration, movement, treatment dose and duration, and other co-occurring conditions that might be influencing the reported medication effects. While we see some consistent medication-related signals across sub-groups (and many ENIGMA studies are the largest of their kind), other factors could still be at play. Additionally, in uncontrolled studies, patients with the most severe symptoms often need the highest doses and experience the greatest brain changes, so medication is confounded with disease severity and duration of illness^62^. Nevertheless, our finding of differential effects on CT compared to SA aligns with randomized clinical trial evidence of medication-related brain structural changes^62^, suggesting that medication status can influence case-control brain profiles across disorders. Further research is warranted to determine the extent to which these findings generalizes to other neuroimaging measures and modalities^62^.

In summary, our study contributes to the ongoing effort to understand the genetic basis of the development of the cerebral cortex and its deviation in psychiatric disorders, by bringing together findings from neuroimaging, psychiatry, and genetics. We reported the distinct neuroimaging profiles observed in NPDs, compared to those associated with individual genetic variants. Our study suggests that the neuroimaging differences in NPDs are likely a manifestation of the aggregated effects of their polygenic architecture, influenced by critical non-genetic factors like medication and the lived experience of the disorder, highlighting the need for integrated models.

## Methods

*Data summary*: In this study, we aggregated multiple datasets as well as published summary statistics from ENIGMA Consortium^31^ to compare case control effect sizes (Cohen’s *d*) across 8 psychiatric disorders and associated common and rare variants on global and regional CT and SA.

*Rare genetic variant participants:* Recurrent deletions and duplications were included in the analysis based on previous analysis of T1-weighted MRI data^41,44,67^, and where at least 18 carriers of the same CNV were available ^41,67^. Clinically ascertained groups: CNV carriers were recruited after either being referred for genetic testing due to the diagnosis of a neurodevelopmental disorder or as the relative (e.g., parent) of a CNV carrier. Controls were defined as individuals who did not carry any NPD-associated CNVs.

Unselected population group: CNV carriers were identified in the UK Biobank. Controls were defined as individuals who did not carry any of the recurrent CNVs selected from this study. Demographic details and coordinates of each of the 11 CNVs are provided in **Supplement Material**. Signed consents were obtained by investigators from each cohort for all participants and/or their legal representatives prior to the investigation. This study, using an aggregate dataset, obtained ethics approval from the CHU Sainte-Justine Hospital.

*Rare genetic variant MRI image acquisition and preprocessing:* The data sample included 3D T1-weighted (T1w) volumetric brain images at 0.8-1 mm isotropic resolution across all sites. MRI parameters for each cohort are detailed in the Supplemental Material. *Quality control:* Visual quality inspection was performed by the two raters (CM, KK) using the ENIGMA standardized quality control protocol (https://github.com/ENIGMA-git).

*NPD participants, MRI image acquisition and preprocessing:* In this study, we aggregated published summary statistics for 8 NPDs from the following published ENIGMA studies: ADHD^49^; ASD^49^; BD ^50^; clinical high-risk for psychosis (CHR-PS)^51^; conduct disorder (CD)^52^; MDD^53^; OCD^49^; and SCZ^55^. All these studies followed the ENIGMA standardized quality control protocol (https://github.com/ENIGMA-git), and processing. An extensive description of MRI image acquisition, methods for pre-processing the data are available in the respective published study.

*Cortical thickness and surface area measures:* FreeSurfer 5.3.0 was used to extract cortical thickness and surface area for 68 Desikan ROIs ^69^, as well as total surface area (total SA), and mean cortical thickness (mean CT). See Supplementary Methods for details.

*Statistical analysis:* Linear regression models (R version 3.6.3) were used to compute CNV-control differences (Cohen’s *d*) for each CNV using age, sex, and site, as covariates. For regional surface area, we additionally used total Surface Area, as a covariate. This approach was used for CNVs. The FDR procedure ^70^ was applied for multiple comparison correction. The significance was set at FDR-corrected *q* < 0.05. See Supplementary Methods for additional details.

*Effect sizes (NPDs and CNVs):* Cohen’s *d* were computed based on case-control linear regression. Cohen’s *d* values for neurodevelopmental and psychiatric disorders were extracted from published ENIGMA studies ^30,31,71^: ADHD ^49^; ASD ^49^; BD ^50^; clinical high-risk for psychosis (CHR-PS)^51^; conduct disorder (CD)^52^; MDD^53^; OCD^54^; and SCZ^55^. All effect sizes were computed after regressing for age, sex, and site. For regional surface area analysis, adjustment for brain size was added either using total SA or ICV as a covariate, except for MDD ^53^, for which such results were unavailable. In addition, we used previously published meta-analysis effect sizes for total SA and mean CT for 9 genetic mutations ^43^ including 6 aneuploidies^37,72,73^ (Turner Syndrome (TS); Down Syndrome; XXX; XXY; XYY; and XXYY) and 3 CNVs (7q11.23^74^, 16p11.2 distal deletion and duplication ^42,75^).

*Effect size comparison metrics:* For comparisons across metrics, the following effect size metrics were used: absolute Cohen’s *d* for global measures, and average absolute Cohen’s *d* of the top decile across regional cortical thickness and surface area ^67^. As the proportion of significant regions varied across CNVs and NPDs (due to differences in effect and sample sizes), we chose to focus on the top decile Cohen’s *d* for all CNVs and NPDs to avoid biases and to provide effect sizes comparable across CNVs and NPDs. Statistical testing of spatially correlated Cohen’s *d* profiles was performed using spin permutation testing ^76,77^. See details in Supplementary Methods. All cortical projections were generated using the *ggseg* R package78.

*Common genetic risk associated with NPDs:* To assess common genetic risk associated with NPDs, we investigated the GWAS summary statistics and genome-wide significant loci from ENIGMA and PGC. Specifically, we leveraged the following published statistics: cortical thickness and surface area^14^; ADHD ^57^; BD^56^; MDD ^58^; and SCZ ^59^. We used the effect sizes from ENIGMA cortical summary statistics^14^ for all the NPD-associated genome-wide significant loci, both global and regional MRI metrics.

*Ranking and significance of NPD-associated genome-wide significant loci:* To investigate whether the common genetic risk factors associated with NPDs are also implicated in the genetic architecture of the human cortex, we conducted the following analysis. We ranked independent single nucleotide polymorphisms (SNPs) from GWAS of NPDs (ADHD, BD, MDD, and SCZ) according to their association with cortical GWAS (cortical thickness and surface area). Subsequently, we calculated the median rank of NPD SNPs within the ranked cortical GWAS. To assess the significance of this observation, we performed 10,000 null permutations by randomly sampling an equal number of SNPs and recomputing the median rank. This yielded a permutation-based p-value for each NPD. Finally, we applied a significance threshold (FDR q < 0.05) based on the permutation testing. This was also assessed for the cortical thickness and surface area SNPs, by ranking them based on NPD GWASes. SNPs from SCZ and cortical thickness GWAS were excluded from further analyses due to the lack of significant enrichment.

*Consensus maps of regional cortical thickness and surface area differences*: We computed consensus maps (per ROI across CNVs/NPDs) using: i) mean absolute effect size; ii) % significance; iii) variance in effect size values; and iv) latent dimensions of cortical differences across CNVs and across NPDs: Principal component analysis identified latent dimensions of cortical thickness and surface area differences across all CNVs and across all NPDs separately. We used the FactoMineR^79^ package in R and ran PCA on Cohen’s *d* values.

*Correlation with normative maps of cortical gradients*: We used the previously published normative maps of cortical organization hierarchies ^39^, specifically, the spatial transcriptomic map (PC1 gene expression) ^38,40^. To compute a spatial correlation with the normative map, we mapped these maps to 68 cortical regions of Desikan parcellation using the neuromaps python package ^39^. Finally, a spin permutation method ^76,77^ was used to assess the significance of the spatial correlation.

*Polygenic Risk Scores (PRS):* We used the standard PRS scores for bipolar disorder and schizophrenia, provided by the UK Biobank (data fields 26214 and 26275)^80^. We only kept individuals of European Ancestry and removed any recurrent CNV carriers.

## Data and materials availability

UK Biobank data was downloaded under the application 40980 and may be accessed via their standard data access procedure (see http://www.ukbiobank.ac.uk/register-apply). UK Biobank CNVs were called using the pipeline developed in the Jacquemont Lab, as described at https://github.com/MartineauJeanLouis/MIND-GENESPARALLELCNV. The final CNV calls are available for download from the UK Biobank returned datasets (Return ID: 3104, https://biobank.ndph.ox.ac.uk/ukb/dset.cgi?id=3104). The 22q11.2 UCLA raw data are currently available by request from the project PI. Raw neuroimaging data for rare variants are available through request and data access agreement from the PIs of the projects (Brain Canada: S.J. CHUSJ Montreal; 22q11.2: C.E.B. UCLA, Cardiff: D.E.J.L., M.J.O., M.V.B., J.H, Cardiff University; SCA: A.R. NIMH). References to the processing pipeline and R package versions used for analysis are listed in the methods.

## Code availability

The code for generating all the figures and supplement figures, along with processed summary measures is available in the following GitHub repository: https://github.com/kkumar-iitkgp/ct_sa_across_disorders_and_variants.git

## Acknowledgments

### Funding

This research was supported by Calcul Quebec (http://www.calculquebec.ca) and Compute Canada (http://www.computecanada.ca), the Brain Canada Multi-Investigator initiative, NIH U01 grant for CAMP (1U01MH119690-01), the Canadian Institutes of Health Research, CIHR_400528, The Institute of Data Valorization (IVADO) through the Canada First Research Excellence Fund, Healthy Brains for Healthy Lives through the Canada First Research Excellence Fund. Dr Jacquemont is a recipient of a Canada Research Chair in neurodevelopmental disorders and a chair from the Jeanne et Jean Louis Levesque Foundation. The Cardiff CNV cohort was supported by the Wellcome Trust Strategic Award “DEFINE” and the National Centre for Mental Health with funds from Health and Care Research Wales (code 100202/Z/12/Z). The CHUV cohort was supported by the SNF (Maillard Anne, Project, PMPDP3 171331). Data from the UCLA cohort provided by Dr. Bearden (participants with 22q11.2 deletions or duplications and controls) was supported through grants from the NIH (U54EB020403), NIMH (R01MH085953, R01MH100900, R03MH105808), and the Simons Foundation (SFARI Explorer Award). Claudia Modenato was supported by the doc.mobility grant provided by the Swiss National Science Foundation (SNSF). Kuldeep Kumar was supported by The Institute of Data Valorization (IVADO) Postdoctoral Fellowship program, through the Canada First Research Excellence Fund. CRKC and PMT are supported in part by NIMH grants R01MH116147, R01MH123163, and R01MH121246, and by the Milken Institute and the Baszucki Brain Research Fund. Dr. Sønderby is supported by the Research Council of Norway (#223273), South-Eastern Norway Regional Health Authority (#2020060), European Union’s Horizon2020 Research and Innovation Programme (CoMorMent project; Grant #847776) and Kristian Gerhard Jebsen Stiftelsen (SKGJ-MED-021). BD is supported by the Swiss National Science Foundation (NCCR Synapsy, project grant numbers 32003B_135679, 32003B_159780, 324730_192755, and CRSK-3_190185), the Roger De Spoelberch and the Leenaards Foundations. G.D. is supported by the Institute for Data Valorization, Montreal (IVADO; CF00137433), the Fonds de recherche du Québec (FRQ; 285289), the Natural Sciences and Engineering Research Council of Canada (NSERC; DGECR-2023-00089), and the Azrieli Global Scholars Fellowship from the Canadian Institute for Advanced Research (CIFAR) in the Brain, Mind, & Consciousness program. We thank all of the families participating at the Simons Searchlight sites, as well as the Simons Searchlight Consortium. We appreciate obtaining access to imaging and phenotypic data on SFARI Base. Approved researchers can obtain the Simons Searchlight population dataset described in this study by applying at https://base.sfari.org. We are grateful to all families who participated in the 16p11.2 European Consortium.

### Disclosures

MvdB reports grants from Takeda Pharmaceuticals, outside the submitted work. P.M.T. and CRKC received a research grant from Biogen, Inc., for work unrelated to this manuscript. All other authors reported no biomedical financial interests or potential conflicts of interest.

### Author contributions

K.K., Z.L., C.Mod., C.C, C.E.B., P.M.T., T.P., and S.J. designed the study, analyzed imaging data, and drafted the manuscript.

*<u>Analyses:</u>* K.K. and C.Mod. performed all the analyses of neuroimaging data. W.S. and A.R. performed analyses of neuroimaging data from sex chromosome aneuploidies.

*<u>Data collection:</u>* C.Mod., A.M., B.R-H., A.P., S.R., and S.M-B. recruited and scanned participants in the 16p11.2 European Consortium. S.L., C.O.M., E.D., F. T-D., V.C., A.R.C.,

F.D. recruited and scanned participants in the Brain Canada cohort. L.K., C.E.B. collected and provided the data for the UCLA cohort. D.E.J.L., M.J.O., M.B.M. V.d.B., J.H., and

A.I.S., provided the data for the Cardiff cohort. W.S. and A.R. provided the data for sex chromosome aneuploidies.

All authors provided feedback on the manuscript.

### Article Information

Each cohort and corresponding study received approval from their local institutional review board and this study was approved by the Institutional review board of the CHU Ste Justine research center. The Simons Searchlight Consortium principal investigator is Wendy K. Chung. Contributors to the Simons Searchlight Consortium include the following: Hanalore Alupay, BS, Benjamin Aaronson, BS, Sean Ackerman, MD, Katy Ankenman, MSW, Ayesha Anwar, BA, Constance Atwell, PhD, Alexandra Bowe, BA, Arthur L. Beaudet, MD, Marta Benedetti, PhD, Jessica Berg, MS, Jeffrey Berman, PhD, Leandra N. Berry, PhD, Audrey L. Bibb, MS, Lisa Blaskey, PhD, Jonathan Brennan, PhD, Christie M. Brewton, BS, Randy Buckner, PhD, Polina Bukshpun, BA, Jordan Burko, BA, Phil Cali, EdS, Bettina Cerban, BA, Yishin Chang, MS, Maxwell Cheong, BE, MS, Vivian Chow, BA, Zili Chu, PhD, Darina Chudnovskaya, BS, Lauren Cornew, PhD, Corby Dale, PhD, John Dell, BS, Allison G. Dempsey, PhD, Trent Deschamps, BS, Rachel Earl, BA, James Edgar, PhD, Jenna Elgin, BS, Jennifer Endre Olson, PsyD, Yolanda L Evans, MA, Anne Findlay, MA, Gerald D Fischbach, MD, Charlie Fisk, BS, Brieana Fregeau, BA, Bill Gaetz, PhD, Leah Gaetz, MSW, BSW, BA, Silvia Garza, BA, Jennifer Gerdts, PhD, Orit Glenn, MD, Sarah E Gobuty, MS, CGC, Rachel Golembski, BS, Marion Greenup, MPH, MEd, Kory Heiken, BA, Katherine Hines, BA, Leighton Hinkley, PhD, Frank I. Jackson, BS, Julian Jenkins III, PhD, Rita J. Jeremy, PhD, Kelly Johnson, PhD, Stephen M. Kanne, PhD, Sudha Kessler, MD, Sarah Y. Khan, BA, Matthew Ku, BS, Emily Kuschner, PhD, Anna L. Laakman, MEd, Peter Lam, BS, Morgan W. Lasala, BA, Hana Lee, MPH, Kevin LaGuerre, MS, Susan Levy, MD, Alyss Lian Cavanagh, MA, Ashlie V. Llorens, BS, Katherine Loftus Campe, MEd, Tracy L. Luks, PhD, Elysa J. Marco, MD, Stephen Martin, BS, Alastair J. Martin, PhD, Gabriela Marzano, HS, Christina Masson, BFA, Kathleen E. McGovern, BS, Rebecca McNally Keehn, PhD, David T. Miller, MD, PhD, Fiona K. Miller, PhD, Timothy J. Moss, MD, PhD, Rebecca Murray, BA, Srikantan S. Nagarajan, PhD, Kerri P. Nowell, MA, Julia Owen, PhD, Andrea M. Paal, MS, Alan Packer, PhD, Patricia Z. Page, MS, Brianna M. Paul, PhD, Alana Peters, BS, Danica Peterson, MPH, Annapurna Poduri, PhD, Nicholas J. Pojman, BS, Ken Porche, MS, Monica B. Proud, MD, Saba Qasmieh, BA, Melissa B. Ramocki, MD, PhD, Beau Reilly, PhD, Timothy P. L. Roberts, PhD, Dennis Shaw, MD, Tuhin Sinha, PhD, Bethanny Smith-Packard, MS, CGC, Anne Snow Gallagher, PhD, Vivek Swarnakar, PhD, Tony Thieu, BA, MS, Christina Triantafallou, PhD, Roger Vaughan, PhD, Mari Wakahiro, MSW, Arianne Wallace, PhD, Tracey Ward, BS, Julia Wenegrat, MA, and Anne Wolken, BS. European 16p11.2 Consortium principal investigator Sébastien Jacquemont. Members of the European 16p11.2 Consortium include the following: Addor Marie-Claude, Service de génétique médicale, Centre Hospitalier Universitaire Vaudois, Lausanne University, Switzerland; Andrieux Joris, Institut de Génétique Médicale, CHRU de Lille, Hopital Jeanne de Flandre, France; Arveiler Benoît, Service de génétique médicale, CHU de Bordeaux- GH Pellegrin, France; Baujat Geneviève, Service de Génétique Médicale, CHU Paris - Hôpital Necker-Enfants Malades, France; Sloan-Béna Frédérique, Service de médecine génétique, Hôpitaux Universitaires de Genève - HUG, Switzerland; Belfiore Marco, Service de génétique médicale, Centre Hospitalier Universitaire Vaudois, Lausanne University, Switzerland; Bonneau Dominique, Service de génétique médicale, CHU d’Angers, France; Bouquillon Sonia, Institut de Génétique Médicale, Hopital Jeanne de Flandre, Lille, France; Boute Odile, Hôpital Jeanne de Flandre, CHRU de Lille, Lille, France; Brusco Alfredo, Genetica Medica, Dipartimento di Scienze Mediche, Università di Torino, Italy; Busa Tiffany, Département de génétique médicale, CHU de Marseille, Hôpital de la Timone, France; Caberg Jean- Hubert, Centre de génétique humaine, CHU de Liège, Belgique; Campion Dominique, Service de psychiatrie, Centre hospitalier de Rouvray, Sotteville lès Rouen, France; Colombert Vanessa, Service de génétique médicale, Centre Hospitalier Bretagne Atlantique CH Chubert- Vannes, France; Cordier Marie-Pierre, Service de génétique clinique, CHU de Lyon, Hospices Civils de Lyon, France; David Albert, Service de Génétique Médicale, CHU de Nantes, Hôtel Dieu, France; Debray François-Guillaume, Service de Génétique Humaine, CHU Sart Tilman - Liège, Belgique; Delrue Marie-Ange, Service de génétique médicale, CHU de Bordeaux, Hôpital Pellegrin, France; Doco-Fenzy Martine, Service de Génétique et Biologie de la Reproduction, CHU de Reims, Hôpital Maison Blanche, France; Dunkhase- Heinl Ulrike, Department of Pediatrics, Aabenraa Hospital, Sonderjylland, Denmark; Edery Patrick, Service de génétique clinique, CHU de Lyon, Hospices Civils de Lyon, France; Fagerberg Christina, Department of Clinical Genetics, Odense University hospital, Denmark; Faivre Laurence, Centre de génétique, Hôpital d’Enfants, CHU Dijon Bourgogne - Hôpital François Mitterrand, France; Forzano Francesca, Ambulatorio di Genetica Medica, Ospedali Galliera di Genova, Italy and Clinical Genetics Department, 7^th^ Floor Borough Wing, Guy’s Hospital, Guy’s & St Thomas’ NHS Foundation Trust, Great Maze Pond, London SE1 9RT, UK; Genevieve David, Département de Génétique Médicale, Maladies Rares et Médecine Personnalisée, service de génétique clinique, Université Montpellier, Unité Inserm U1183, CHU Montpellier, Montpellier, France; Gérard Marion, Service de Génétique, CHU de Caen, Hôpital Clémenceau, France; Giachino Daniela, Genetica Medica, Dipartimento di Scienze Cliniche e Biologiche, Università di Torino, Italy; Guichet Agnès, Service de génétique, CHU d’Angers, France; Guillin Olivier, Service de psychiatrie, Centre hospitalier du Rouvray, Sotteville lès Rouen, France; Héron Delphine, Service de Génétique clinique, CHU Paris-GH La Pitié Salpêtrière-Charles Foix - Hôpital Pitié Salpêtrière, France; Isidor Bertrand, Service de Génétique Médicale, CHU de Nantes, Hôtel Dieu, France; Jacquette Aurélia, Service de Génétique clinique, CHU Paris-GH La Pitié Salpêtrière-Charles Foix - Hôpital Pitié-Salpêtrière, France; Jaillard Sylvie, Service de Génétique Moléculaire et Génomique – Pôle biologie, CHU de Rennes, Hôpital Pontchaillou, France; Journel Hubert, Service de génétique médicale, Centre Hospitalier Bretagne Atlantique CH Chubert- Vannes, France; Keren Boris, Centre de Génétique Moléculaire et Chromosomique, CHU Paris-GH La Pitié Salpêtrière-Charles Foix - Hôpital Pitié-Salpêtrière, France; Lacombe Didier, Service de génétique médicale, CHU de Bordeaux-GH Pellegrin, France; Lebon Sébastien, Pediatric Neurology Unit, Department of Pediatrics, Lausanne University Hospital, Lausanne, Switzerland; Le Caignec Cédric, Service de Génétique Médicale - Institut de Biologie, CHU de Nantes, France; Lemaître Marie-Pierre, Service de Neuropédiatrie, Centre Hospitalier Régional Universitaire de Lille, France; Lespinasse James, Service génétique médicale et oncogénétique, Hotel Dieu, Chambéry, France; Mathieu-Dramart Michèle, Service de Génétique Clinique, CHU Amiens Picardie, France; Mercier Sandra, Service de Génétique Médicale, CHU de Nantes, Hôtel Dieu, France; Mignot Cyril, Service de Génétique clinique, CHU Paris-GH La Pitié Salpêtrière-Charles Foix - Hôpital Pitié-Salpêtrière, France; Missirian Chantal, Département de génétique médicale, CHU de Marseille, Hôpital de la Timone, France; Petit Florence, Service de génétique clinique Guy Fontaine, Hôpital Jeanne de Flandre, CHRU de Lille, France; Pilekær Sørensen Kristina, Department of Clinical Genetics, Odense University Hospital, Denmark; Pinson Lucile, Département de Génétique Médicale, Maladies Rares et Médecine Personnalisée, service de génétique clinique, Université Montpellier, Unité Inserm U1183, CHU Montpellier, Montpellier, France; Plessis Ghislaine, Service de Génétique, CHU de Caen, Hôpital Clémenceau, France; Prieur Fabienne, Service de génétique clinique, CHU de Saint-Etienne - Hôpital Nord, France; Raymond Alexandre, Center for Integrative Genomics, Lausanne University, Switzerland; Rooryck-Thambo Caroline, Laboratoire de génétique moléculaire, CHU de Bordeaux-GH Pellegrin, France; Rossi Massimiliano, Service de génétique clinique, CHU de Lyon, Hospices Civils de Lyon, France; Sanlaville Damien, Laboratoire de Cytogénétique Constitutionnelle, CHU de Lyon, Hospices Civils de Lyon, France; Schlott Kristiansen Britta, Department of Clinical Genetics, Odense University Hospital, Denmark; Schluth-Bolard Caroline, Laboratoire de Cytogénétique Constitutionnelle, CHU de Lyon, Hospices Civils de Lyon, France; Till Marianne, Service de génétique clinique, CHU de Lyon, Hospices Civils de Lyon, France; Van Haelst Mieke, Department of Genetics, University Medical Center Utrecht, Holland; Van Maldergem Lionel, Centre de Génétique humaine, CHRU de Besançon - Hôpital Saint-Jacques, France.

