## Supplement File for "Cortical differences across psychiatric disorders and associated common and rare genetic variants"

### Supplement Material

### MRI data acquisition and quality check

##### Participants

Clinically ascertained CNV carriers were recruited as either probands referred for genetic testing or as relatives. Controls were either non-carriers within the same families or individuals from the general population. We pooled data from 5 different cohorts: Cardiff University (Cardiff, UK), 16p11.2 European Consortium (Lausanne, Switzerland), University of Montreal (Canada), UCLA (Los Angeles, USA), and the Variation in Individuals Project (SVIP, USA). A subset of the participants with 16p11.2 proximal and 22q11.2 CNVs were included in prior publications [1–4]. CNVs from non-clinical populations were identified in the UK Biobank [5,6]. PennCNV and QuantiSNP were used, with standard quality control metrics, to identify CNVs [7–9].

##### 16p11.2 European Consortium

MRI data of the EU participants were acquired on two 3T whole-body scanners. 14 carriers of a 16p11.2 proximal deletion and 17 duplication carriers, together with 59 controls (21 familial and 38 unrelated controls) were examined on a Magnetom TIM Trio (Siemens Healthcare, Erlangen, Germany), using a 12-channel RF receive head coil and RF body transmit coil. The remaining 16p11.2 proximal (13 deletions, 6 duplications), 1q21.2 distal (9 deletions, 7 duplications) carriers and controls (n=38) in the European cohort, were scanned on a Magnetom Prisma Syngo (Siemens Healthcare, Erlangen, Germany) using a 64-channel RF receive head coil and RF body transmit coil. T1-weighted (T1w) anatomical images acquired with the TIM Trio scanner used a Multi-Echo Magnetization Prepared RApid Gradient Echo sequence (ME-MPRAGE: 176 slices; 256×256 matrix; echo time (TE): TE1 = 1.64 ms, TE2 = 3.5 ms, TE3 = 5.36 ms, TE4 = 7.22 ms; repetition time (TR):

2530 ms; flip angle 7°). On the Prisma Syngo scanner, T1w images were acquired using a single-echo MPRAGE sequence (176 slices; 256×256 matrix; TE = 2.39 ms; TR = 2000 ms; flip angle 9°).

##### Simons Searchlight Consortium

Data were acquired using multi and single-echo sequences. 176 participants (38 del/ 34 dup 16p11.2 proximal carriers, 2 dup 1q21.1 distal carriers, and 102 familial controls) underwent the research MRI protocol at two imaging core sites on matched 3T Magnetom TIM Trio MRI scanners (Siemens Healthcare, Erlangen, Germany), using the vendor-supplied 32-channel phased-array radio-frequency head coils. 68 participants were scanned at University of California sites (UC) and 108 at the Children's Hospital of Philadelphia (CHOP). Structural MRI data included multi-echo T1w ME-MPRAGE using the following parameters: 176 slices, 256×256 matrix, TR = 2530 ms, TI = 1200 ms, TE = 1.64 ms, and flip angle 7°. Clinical MRI images (single-echo) obtained at the phenotyping core sites were also analyzed. The remaining 79 subjects (19 del/ 13 dup 16p11.2 proximal carriers, 12 del/8 dup 1q21.1 distal carriers and 27 familial controls) were scanned at the University of Washington Medical Center, Baylor University Medical Center, and Boston Children's Hospital on two matched 3T Philips Achieva (Philips Healthcare, United States of America) and one unmatched Magnetom TIM Trio scanner (Siemens Healthcare, Erlangen, Germany), respectively. T1w images were acquired using a single-echo MPRAGE sequence and the following parameters: 160 slices; 256×256 matrix; TE = 2.98 ms; TR = 2300 ms; flip angle 9°. All multi-echo images were averaged following a Root-Mean Square (RMS) averaging method.

##### Brain Canada (BC, University of Montreal)

MRI scans for the Brain Canada cohort have been performed at the Montreal Neurological Institute with the same 3T scanner: Magnetom Prisma Syngo (Siemens Healthcare, Erlangen, Germany). Data included 16p11.2 proximal (3 deletions, 3 duplications), 1q21.2 distal (5 deletions, 1 duplication), 22q11.2 (1 duplication) carriers, and controls (n=26) T1w images were acquired using MPRAGE sequences, scanning protocol description is detailed on this website: http://www.bic.mni.mcgill.ca/users/jlewis/BrainCanada/MCIN/.

#### UCLA

Imaging data of 22q11.2 CNV carriers and typically developing (TD) controls were acquired at the University of California, Los Angeles (UCLA). Patients were ascertained from UCLA or Children's Hospital, Los Angeles Pediatric Genetics, Allergy/Immunology, and/or Craniofacial Clinics. We excluded 11 individuals from the analysis due to the insufficient quality of the imaging data (cf. Supplementary Methods, quality control). The final 22q11.2 sample includes 144 individuals (71 deletions, 19 duplications, and 54 controls). Demographically comparable TD comparison subjects were recruited from the same communities as patients via web-based advertisements and by posting flyers and brochures at local schools, pediatric clinics, and other community sites. Exclusion criteria for all study participants included significant neurological or medical conditions (unrelated to 22q11.2 mutation) that might affect brain structure, history of head injury with loss of consciousness, insufficient fluency in English, and/or substance or alcohol abuse or dependence within the past 6 months. The UCLA Institutional Review Board approved all study procedures and informed consent documents. Scanning was conducted on an identical 3 Tesla Siemens Trio MRI scanner with a 12-channel head coil at the University of California at Los Angeles Brain Mapping Center or at the Center for Cognitive Neuroscience.

#### Cardiff

Imaging acquisition in Cardiff was performed on a 3 T General Electric HDx MRI system (GE Medical Systems, Milwaukee, WI) using an eight-channel receive-only head RF coil. T1-weighted structural images were acquired with a 3D fast spoiled gradient echo (FSPGR) sequence (TR = 7.8 ms, TE = 3.0 ms, voxel size = 1 $mm^3$ isomorphic). Data included 1 16p11.2 proximal deletion, 1q21.2 distal (3 deletions, 1 duplication), 22q11.2 (3 deletions, 2 duplications) carriers and 15 controls.

#### MRI quality control

All MRI T1w NIfTI images were visually inspected by the same rater (CM) for head coverage, ghosting, and susceptibility artifacts. Images were also screened after segmentation to ensure good tissue classification accuracy. From the clinically ascertained dataset, 55 subjects were excluded for insufficient image quality or artifacts while from the non-clinically ascertained dataset 52 subjects were excluded following the same criteria. Quality assurance protocol for Freesurfer based cortical reconstructions led to the exclusion of an additional 34 scans. Numbers reported in Table 1 are after exclusion.

#### List of abbreviations

CNV: copy number variants; DEL: deletion; DUP: duplication; NPD: neurodevelopmental and psychiatric disorders; Corr: Pearson correlation; ASD: autism spectrum disorder; ADHD: attention deficit hyperactivity disorder; BD: bipolar disorder; MDD: major depressive disorder; OCD: obsessive-compulsive disorder; SCZ: schizophrenia; PC: principal component; L: left hemisphere; Dim: dimension; ICV: Intracranial Volume; PCA: Principal components analysis.

### Supplement Tables

| CLINICAL ASCERTAINMENT | | | | | | | | | | |
|---|---|---|---|---|---|---|---|---|---|---|
| CNV loci | Copy number | Cohort | Age mean(SD) | Age mean(SD) | Male/Female | Male/Female | TIV mean(SD) | TIV mean(SD) | FSIQ mean(SD) | FSIQ mean(SD) |
| 1q21.1 | Deletions *N=28* | EU *N=9* | 29.43 (18.40) | 18.91 (12.91) | 15/13 | 8/1 | 1.22 (0.14) | 1.15 (0.09) | 90.85 (21.75) *N=25* | 81.67 (21.57) *N=9* |
| | | VIP *N=11* | | 35.11 (20.66) | | 3/8 | | 1.27 (0.15) | | 99.33 (23.96) *N=11* |
| | | BC *N=5* | | 28.25 (18.46) | | 4/1 | | 1.17 (0.17) | | 87 (4.76) *N=5* |
| | | Cardiff *N=3* | | 40.18 (13.15) | | 0/3 | | 1.36 (0.02) | | (-) |
| | Duplications *N=17* | EU *N=6* | 34.29 (17.19) | 37.32 (19.58) | 9/8 | 4/2 | 1.57 (0.11) | 1.58 (0.07) | 95.56 (23.19) *N=16* | 96.57 (11.59) *N=6* |
| | | VIP *N=9* | | 34.25 (15.84) | | 5/4 | | 1.54 (0.13) | | 93.80 (30.17) *N=9* |
| | | BC *N=1* | | 8.34 (-) | | 0/1 | | 1.57 (-) | | 106 (-) *N=1* |
| | | Cardiff *N=1* | | 39.45 (-) | | 0/1 | | 1.76 (-) | | (-) |
| 16p11.2 | Deletions *N=78* | EU *N=24* | 17.14 (11.97) | 21.23 (13.51) | 34/44 | 12/12 | 1.54 (0.17) | 1.44 (0.14) | 82.17 (14.99) *N=64* | 74.38 (14.61) *N=13* |
| | | VIP *N=50* | | 13.82 (9.47) | | 20/30 | | 1.58 (0.17) | | 83.98 (14.36) *N=48* |
| | | BC *N=3* | | 31.64 (10.26) | | 1/2 | | 1.46 (0.09) | | 87 (21) *N=3* |
| | | Cardiff *N=1* | | 43.04 (-) | | 1/0 | | 2.01 (-) | | (-) |
| | Duplications *N=68* | EU *N=21* | 31.01(14.91) | 32.55 (13.20) | 38/30 | 11/10 | 1.33 (0.17) | 1.36 (0.16) | 85 (19.70) *N=63* | 72.71 (16.30) *N=17* |
| | | VIP *N=44* | | 30.52 (15.30) | | 24/20 | | 1.30 (0.15) | | 90.44 (18.49) *N=43* |
| | | BC *N=3* | | 26.89 (25.29) | | 3/0 | | 1.49 (0.42) | | 86.67 (23.50) *N=3* |
| 22q11.2 | Deletions *N=68* | UCLA *N=65* | 16.35 (8.56) | 15.66 (7.24) | 33/35 | 32/33 | 1.30 (0.15) | 1.30 (0.15) | 77.41 (13.51) *N=48* | 77.41 (13.51) *N=48* |
| | | Cardiff *N=3* | | 32.56 (20.60) | | 1/2 | | 1.37 (0.11) | | (-) |
| | Duplications *N=19* | UCLA *N=16* | 19.66 (14.24) | 17.32 (12.51) | 8/11 | 7/9 | 1.47 (0.16) | 1.45 (0.17) | 97.83 (20.34) *N=12* | 99.18 (20.76) *N=11* |
| | | BC *N=1* | | 13.67 (-) | | 0/1 | | 1.50 (-) | | 83 (-) *N=1* |
| | | Cardiff *N=2* | | 44.94 (4.77) | | 1/1 | | 1.55 (0.16) | | (-) |
| Controls *N=317* | | EU *N=96* | 25.91 (14.57) | 30.33 (12.95) | 182/135 | 62/34 | 1.46 (0.15) | 1.49 (0.14) | 106.73 (15.03) *N=224* | 102.15 (12.98) *N=59* |
| | | VIP *N=127* | | 24.03 (14.56) | | 70/57 | | 1.44 (0.14) | | 108.88 (11.82) *N=90* |
| | | UCLA *N=46* | | 13.37 (4.96) | | 26/20 | | 1.39 (0.13) | | 112.43 (20.95) *N=44* |
| | | BC *N=33* | | 33.56 (14.90) | | 17/16 | | 1.52 (0.17) | | 101.11 (13.18) *N=31* |
| | | Cardiff *N=15* | | 40.18 (11.12) | | 7/8 | | 1.54 (0.15) | | (-) |

| NON-CLINICAL ASCERTAINMENT | | | | | | |
|---|---|---|---|---|---|---|
| CNV loci | Copy number | Cohort | Age mean(SD) | Male/Female | TIV mean(SD) | UKB FI mean(SD) |
| 1q21.1 | Deletions *N=12* | UKBB | 59.11 (6.71) | 7/5 | 1.35 (0.12) | -0.8 (0.5) *N=10* |
| | Duplications *N=13* | | 60.56 (7) | 4/9 | 1.55 (0.14) | 0.2 (1.3) *N=11* |
| TAR | Duplications *N=31* | | 60 (8) | 14/17 | 1.48 (0.16) | -0.1 (1.2) *N=27* |
| 13q12.12 | Duplications *N=21* | | 62 (8) | 11/10 | 1.54 (0.15) | 0.1 (1.2) *N=18* |
| 15q11.2 | Deletions *N=108* | | 65 (7.10) | 59/49 | 1.54 (0.15) | -0.3 (0.9) N=99 |
| | Duplications *N=144* | | 64 (7.31) | 77/67 | 1.49 (0.15) | 0 (1.1) *N=132* |
| 16p11.2 | Deletions *N=4* | | 65.6 (3.2) | 3/1 | 1.56 (0.13) | 0.8 (0.5) *N=2* |
| | Duplications *N=7* | | 69.3 (2.1) | 2/5 | 1.29 (0.11) | -1.6 (0.2) *N=5* |
| 16p13.11 | Duplications *N=50* | | 66 (6) | 26/24 | 1.49 (0.17) | -0.2 (1.2) *N=46* |
| 22q11.2 | Duplications *N=7* | | 62 (9.5) | 3/4 | 1.55 (0.17) | -0.2 (1.1) *N=6* |
| Controls *N=465* | | | 62.13 (7.40) | 205/260 | 1.51 (0.16) | 0 (1) *N=445* |

#### Supplement Table 1: Detailed demographics

Legend: Detailed demographics and cohort information. EU: 16p11.2 European Consortium, VIP: Simons Searchlight Consortium, BC: Brain Canada, CNV: Copy Number Variant, SD: Standard deviation, TIV: total intracranial volume, FSIQ: Full-scale IQ, UKB FI: UK Biobank fluid intelligence. CNV carriers and controls from the clinically ascertained group come from 5 different cohorts, while non-clinically ascertained participants were identified in the UK Biobank. UK Biobank fluid intelligence scores (UKB field:20016) were adjusted for age, sex, site, and then z-scored. Part of the data was previously published in [2,11] and [1,12].

### Supplement Figures

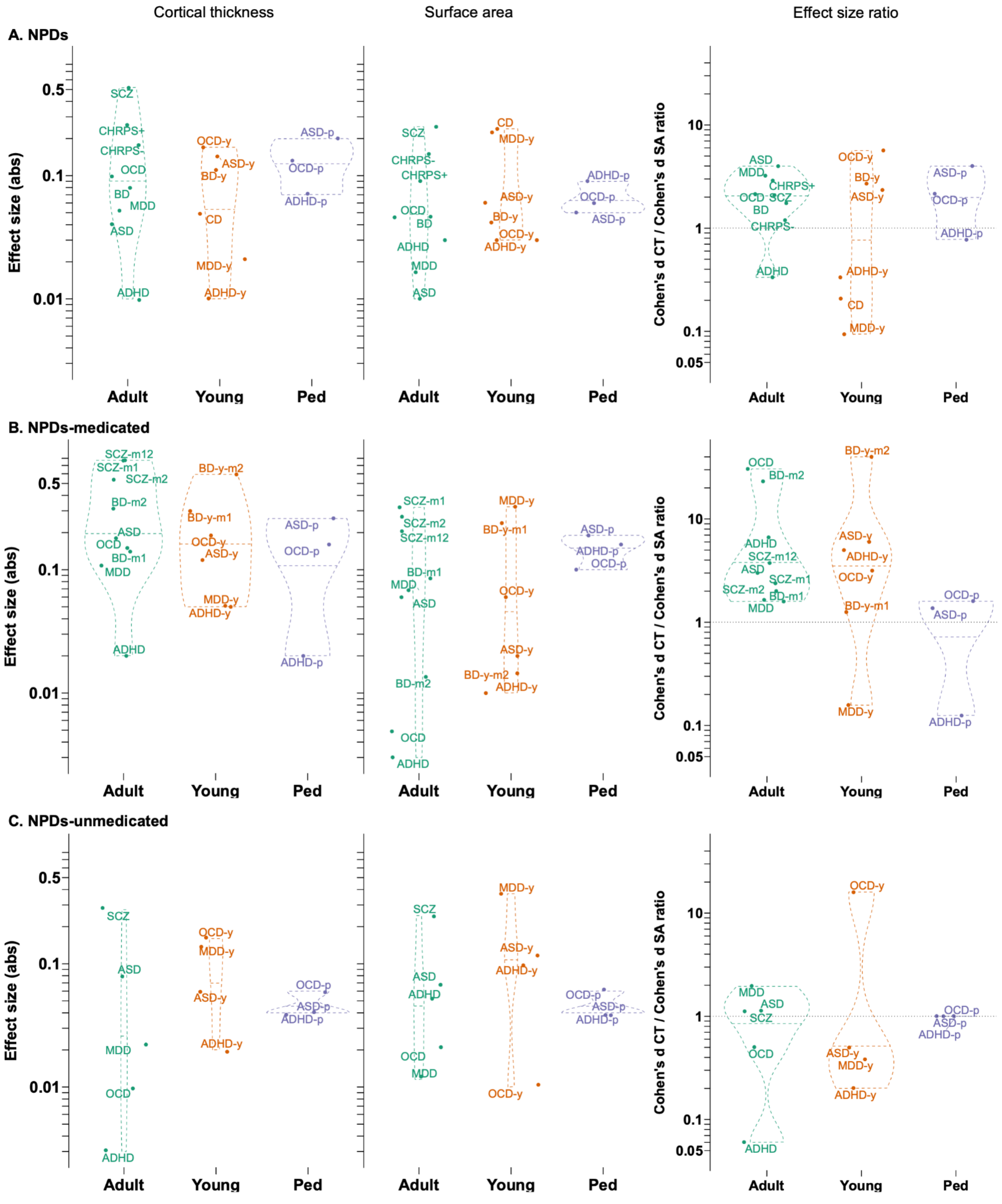


### Supplement Figure 1. Adult-young-pediatric age groups

Legend: Violin plots comparing the absolute effect sizes for mean CT, total SA, and CT/SA effect size ratios for NPDs (top row), NPDs-medicated (middle row), and NPDs-un-medicated (bottom row);

Abbreviations, Abs=absolute; ADHD=attention deficit hyperactivity disorder; ASD=autism spectrum disorder; BD=bipolar disorder; CD: conduct disorder; CHRPS: clinical high risk for psychosis; CNV=copy number variant; CT=cortical thickness; Del=deletion; Dup=duplication; MDD=major depressive disorder; NPD=neurodevelopmental and psychiatric disorders; OCD=obsessive-compulsive disorder; SA=surface area; SCZ=schizophrenia; TS=Turner syndrome;

Adult-Adolescence-Pediatric abbreviations: -y=adolescence/young; -p=pediatric).

Medication abbreviations: -m=medicated; -u=unmedicated; BD-m1=lithium medication; BD-m2=antiepileptics medication; SCZ-m1=1st generation; SCZ-m2=2nd generation; SCZ-m12=1st & 2nd generation;

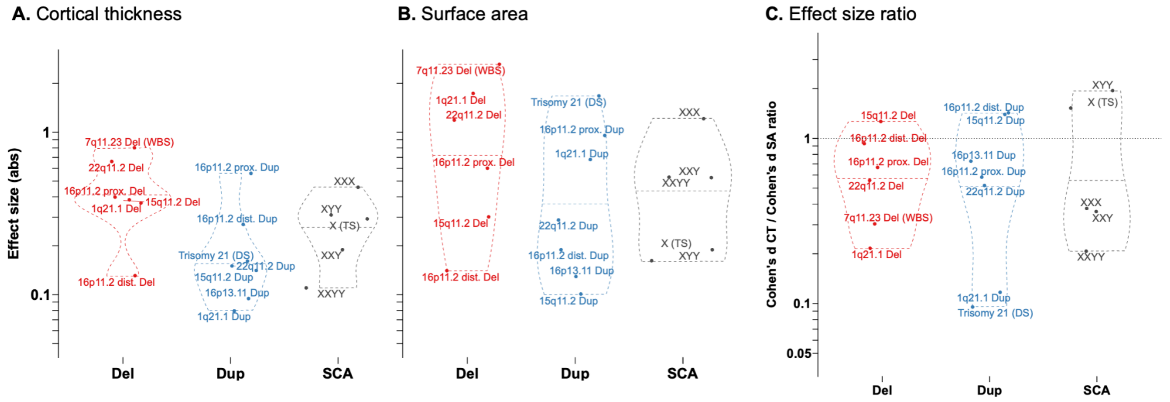


### Supplement Figure 2. Deletion-Duplication-SCA stratifications

Legend: Violin plots comparing the absolute effect sizes for A) mean CT and B) total SA; for deletions (Del), duplications (Dup), sex chromosome aneuploidies (SCA); and C) CT/SA effect size ratios.

Abbreviations, Abs=absolute; ADHD=attention deficit hyperactivity disorder; ASD=autism spectrum disorder; BD=bipolar disorder; CD: conduct disorder; CHRPS: clinical high risk for psychosis; CNV=copy number variant; CT=cortical thickness; Del=deletion; Dup=duplication; MDD=major depressive disorder; NPD=neurodevelopmental and psychiatric disorders; OCD=obsessive-compulsive disorder; SA=surface area; SCZ=schizophrenia; TS=Turner syndrome;

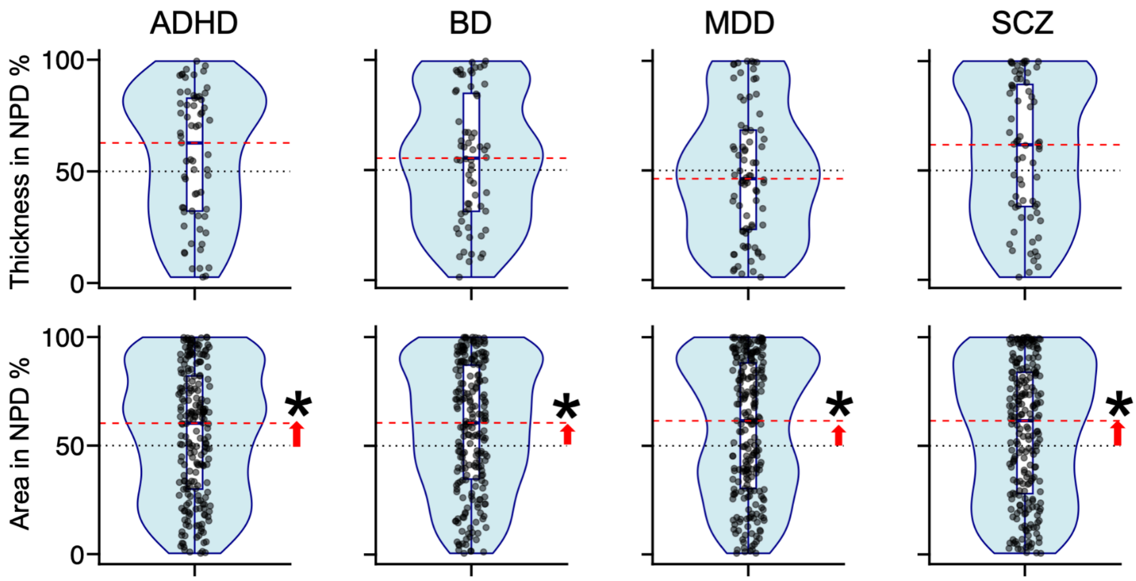


#### Supplement Figure 3. Ranking of cortex-associated genome-wide significant loci in NPD GWAS

Percentiles (%, Y-axis) cortical thickness and area genome-wide significant loci in neurodevelopmental and psychiatric disorders GWAS. Red line: median rank; *: FDR q<0.05; ADHD: attention deficit hyperactivity disorder; BD: bipolar disorder; MDD: major depressive disorder; SCZ: schizophrenia.

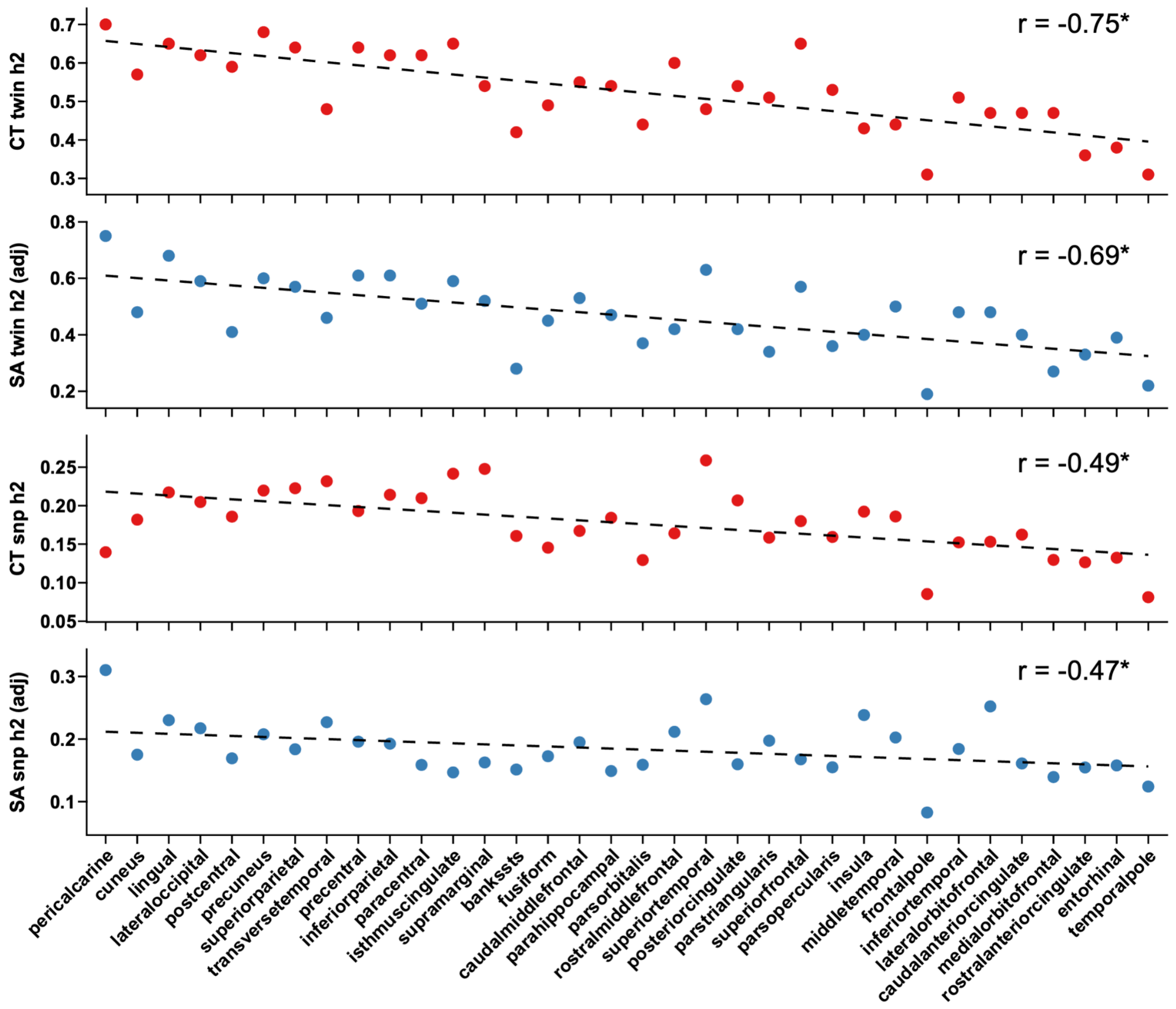


#### Supplement Figure 4. Correlation between regional twin and SNP heritability estimates and cortical gradient.

Legend: Heritability estimates from ENIGMA[13] for cortical thickness (CT) and surface area (SA) across 34 Desikan cortical regions. Regional SA estimates are adjusted for total SA. Y-axis: h2 (twin or SNP heritability), X-axis: cortical regions ordered according to the cortical gradient from sensorimotor to association. Doted line: correlation with the cortical gradient. *: spin permutation p-value < 0.05.

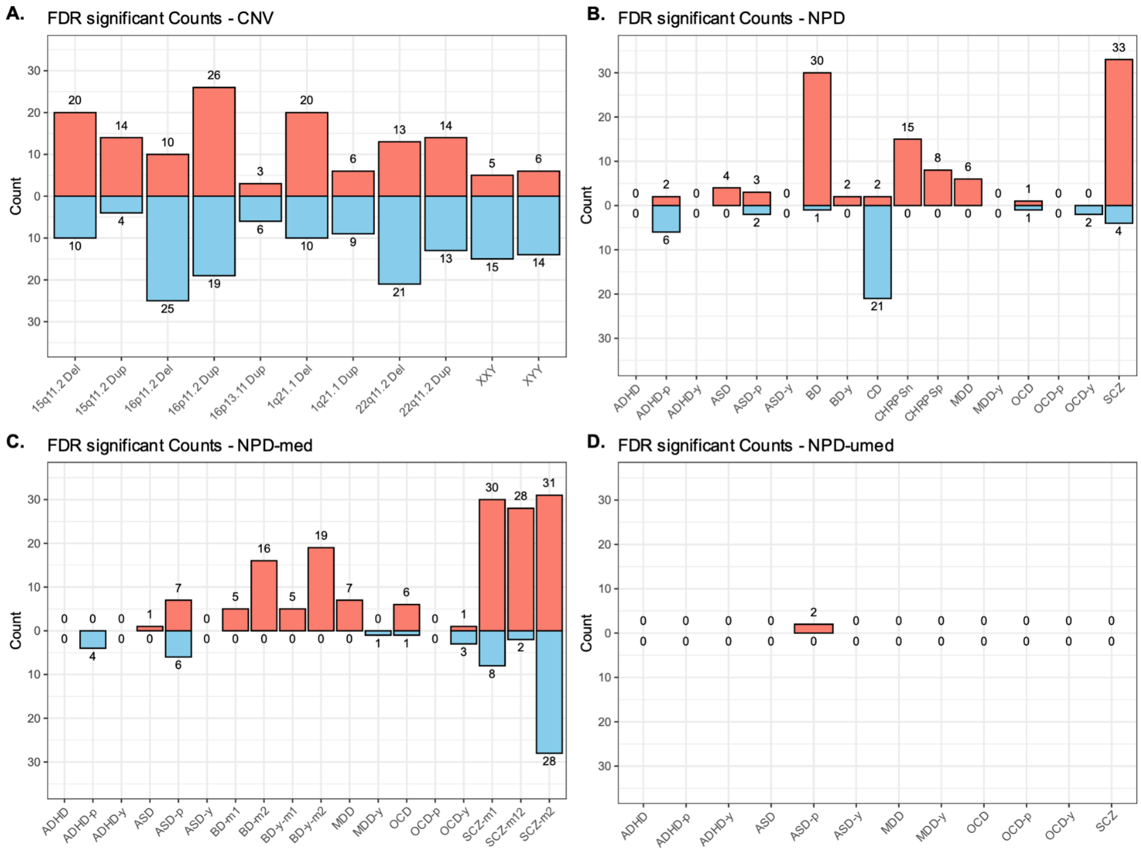


**Supplement Figure 5. Number of FDR significant regions per cortical map**

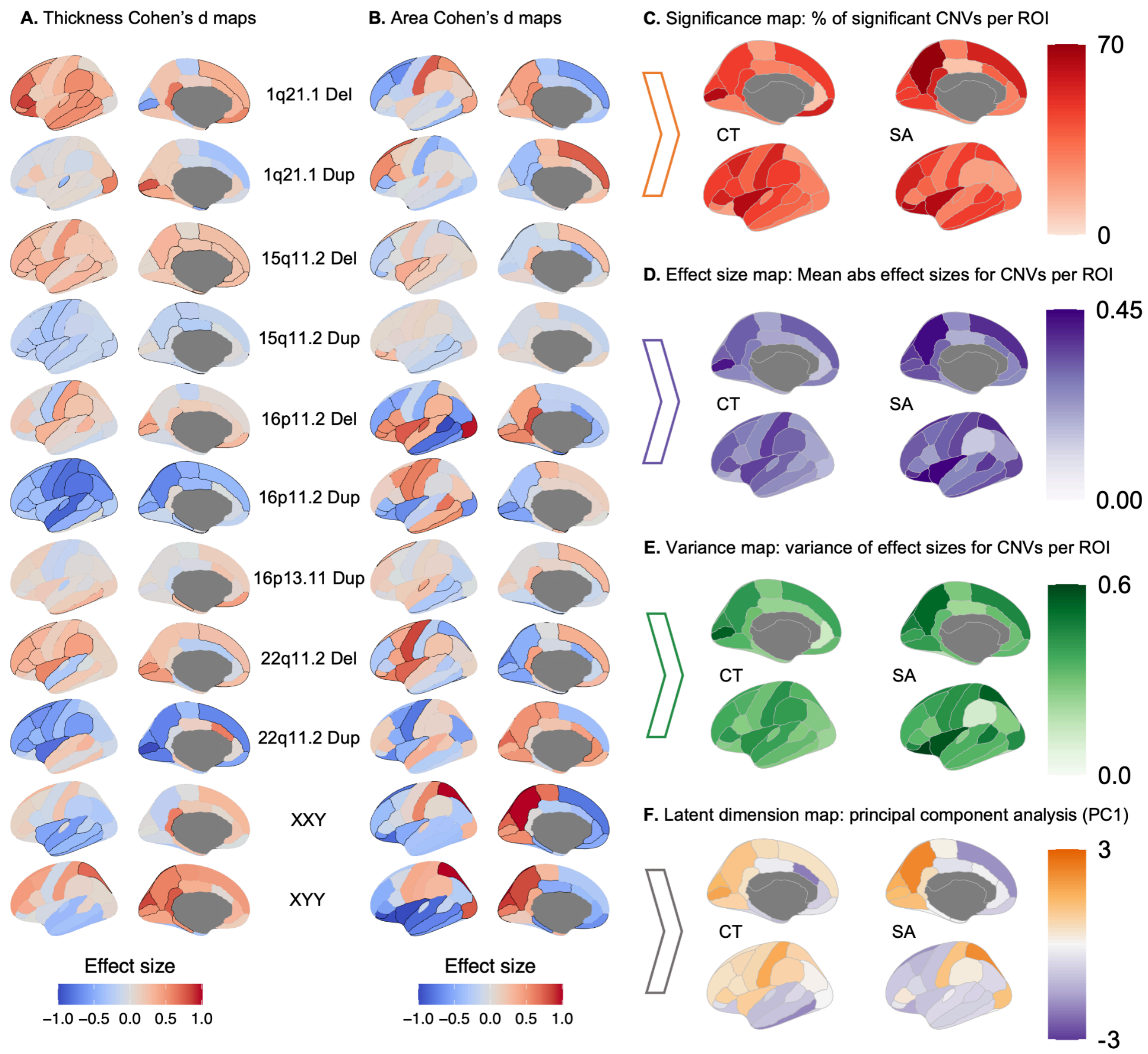


### Supplement Figure 6. Regional cortical alterations and consensus maps across CNVs.

Legend: Cohen's *d* maps for regional cortical alterations for 11 rare genetic risks (CNVs, including aneuploidies): **A**) cortical thickness (CT); **B**) surface area (SA). Case-control differences were calculated after adjusting for age, sex, and site (and total SA for SA). FDR ($q<0.05$) significant regions are shown in black boundaries. **C-F**) Consensus summary maps across CNVs, showing **C**) Significance maps: the percentage of significant alterations per cortical region across CNVs for CT and SA; **D**) effect size maps: the mean of absolute effect sizes per cortical region across CNVs for CT and SA; **E**) variance maps: the variance of effect sizes per cortical region across CNVs for CT and SA; and **F**) Latent dimension map: the first principal component from the principal component analysis (PCA) across CT and SA alterations across CNVs.

Abbreviations. CNV=copy number variants; Corr.=correlation; CT: cortical thickness; Del=deletions; Dup=duplications; SA=surface area; SCZ=schizophrenia; SSD=schizophrenia spectrum disorder;

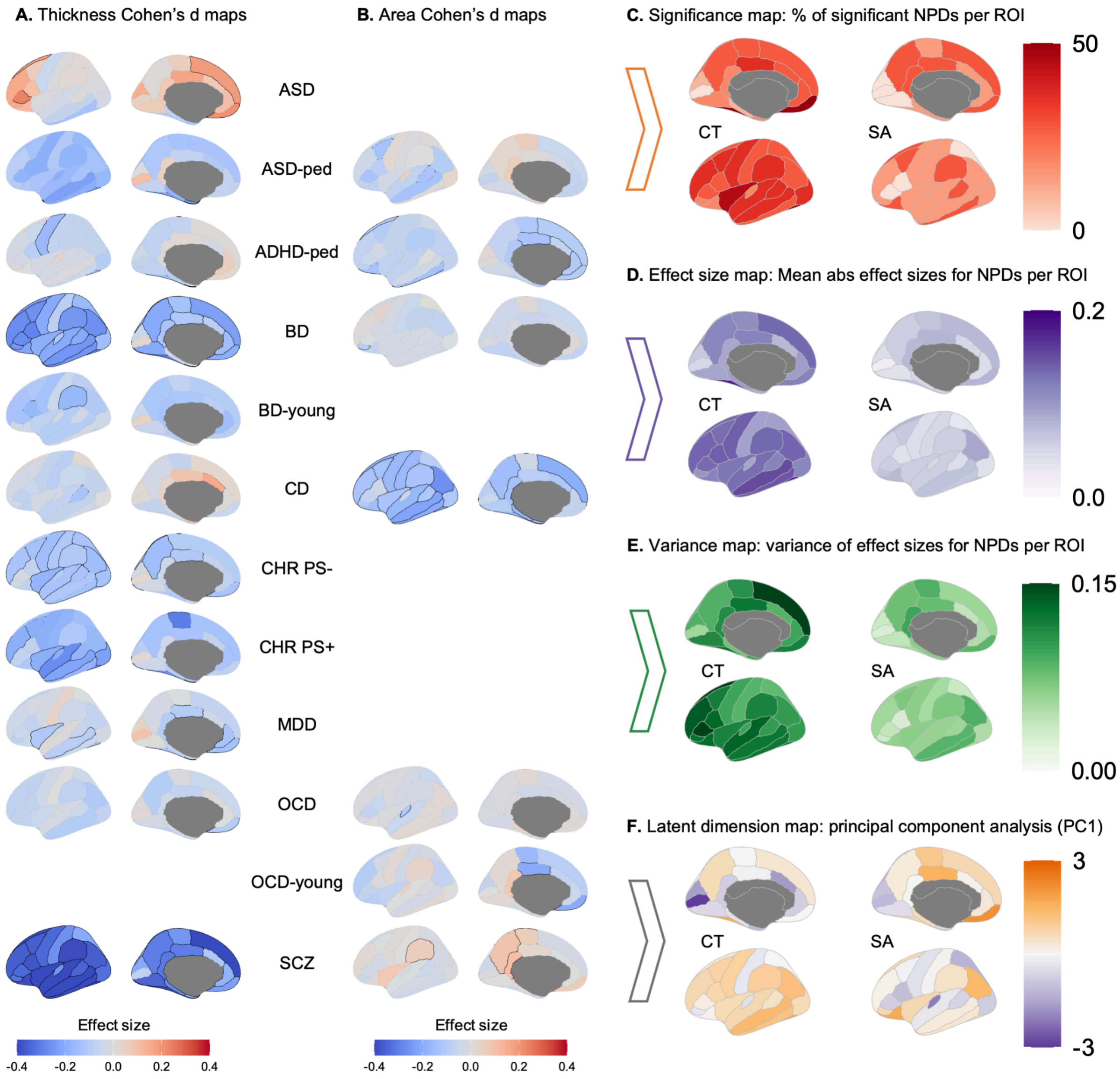


### Supplement Figure 7. Regional cortical alterations and consensus maps across NPDs.

Legend: Cohen's *d* maps for regional cortical alterations for NPDs: **A**) cortical thickness (CT); **B**) surface area (SA). Case-control differences were calculated after adjusting for age, sex, and site (and ICV for SA). FDR ($q<0.05$) significant regions are shown in black boundaries. Only profiles with at least one FDR significant ROI are shown. **C-F**) Consensus summary maps across NPDs, showing **C**) Significance maps: the percentage of significant alterations per cortical region across NPDs for CT and SA; **D**) effect size maps: the mean of absolute effect sizes per cortical region across NPDs for CT and SA; **E**) variance maps: the variance of effect sizes per cortical region across NPDs for CT and SA; and **F**) Latent dimension map: the first principal component from the principal component analysis (PCA) across CT and SA alterations across NPDs.

Abbreviations. ADHD=attention deficit hyperactivity disorder; ASD=autism spectrum disorder; BD=bipolar disorder; Corr.=correlation; CT: cortical thickness; MDD=major depressive disorder; NPDs=neurodevelopmental and psychiatric disorders; OCD=obsessive-compulsive disorder; SA=surface area; SCZ=schizophrenia; SSD=schizophrenia spectrum disorder;

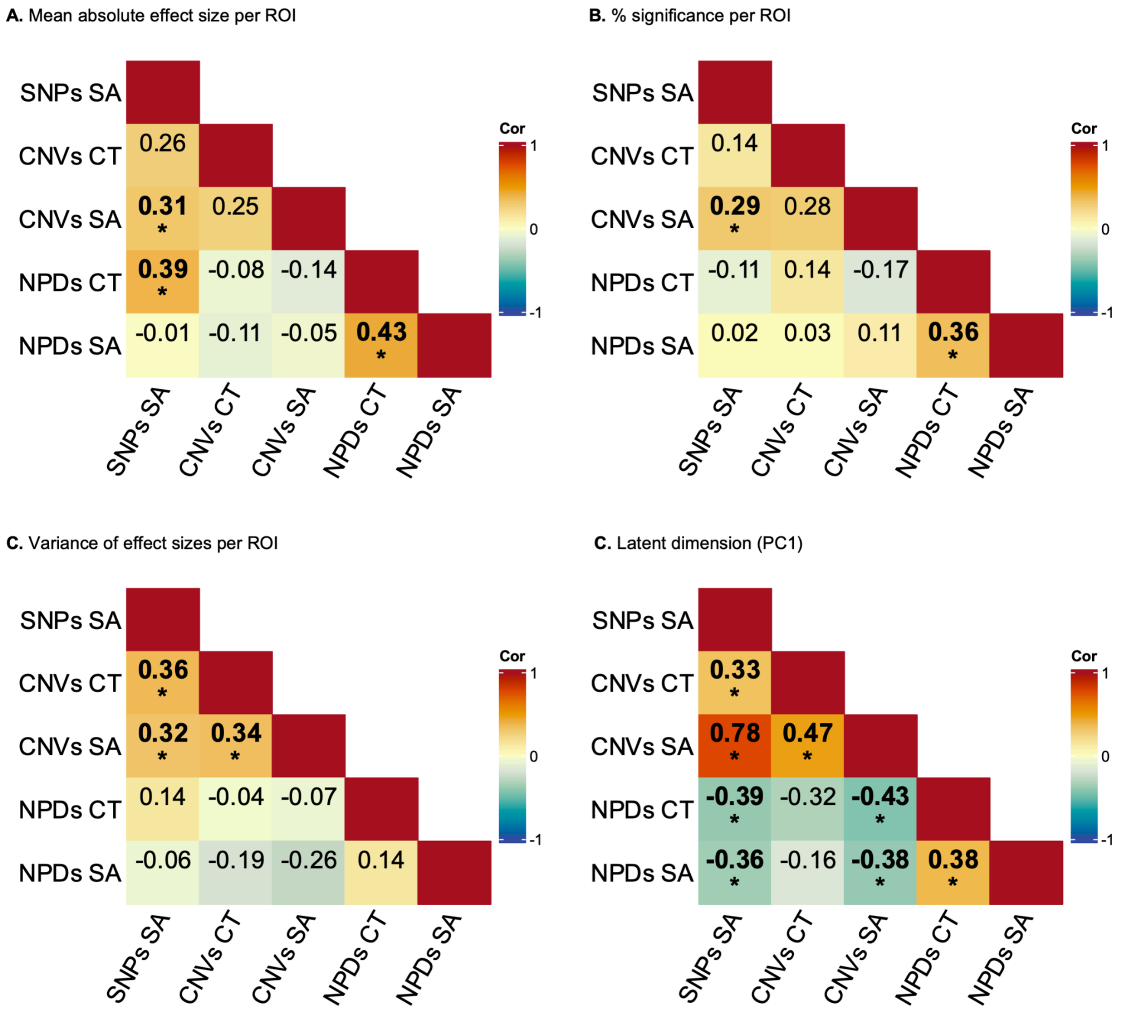


### Supplement Figure 8. Correlations between consensus maps

Legend: Correlation between pairs of consensus maps for **A)** mean absolute effect sizes, **B)** % significance, **C**) variance of effect sizes, and **D**) principal component 1 (PC1). *: spin permutation p-value < 0.05.

Abbreviations. CNV=copy number variants; Cor.=correlation; CT: cortical thickness; SA=surface area;

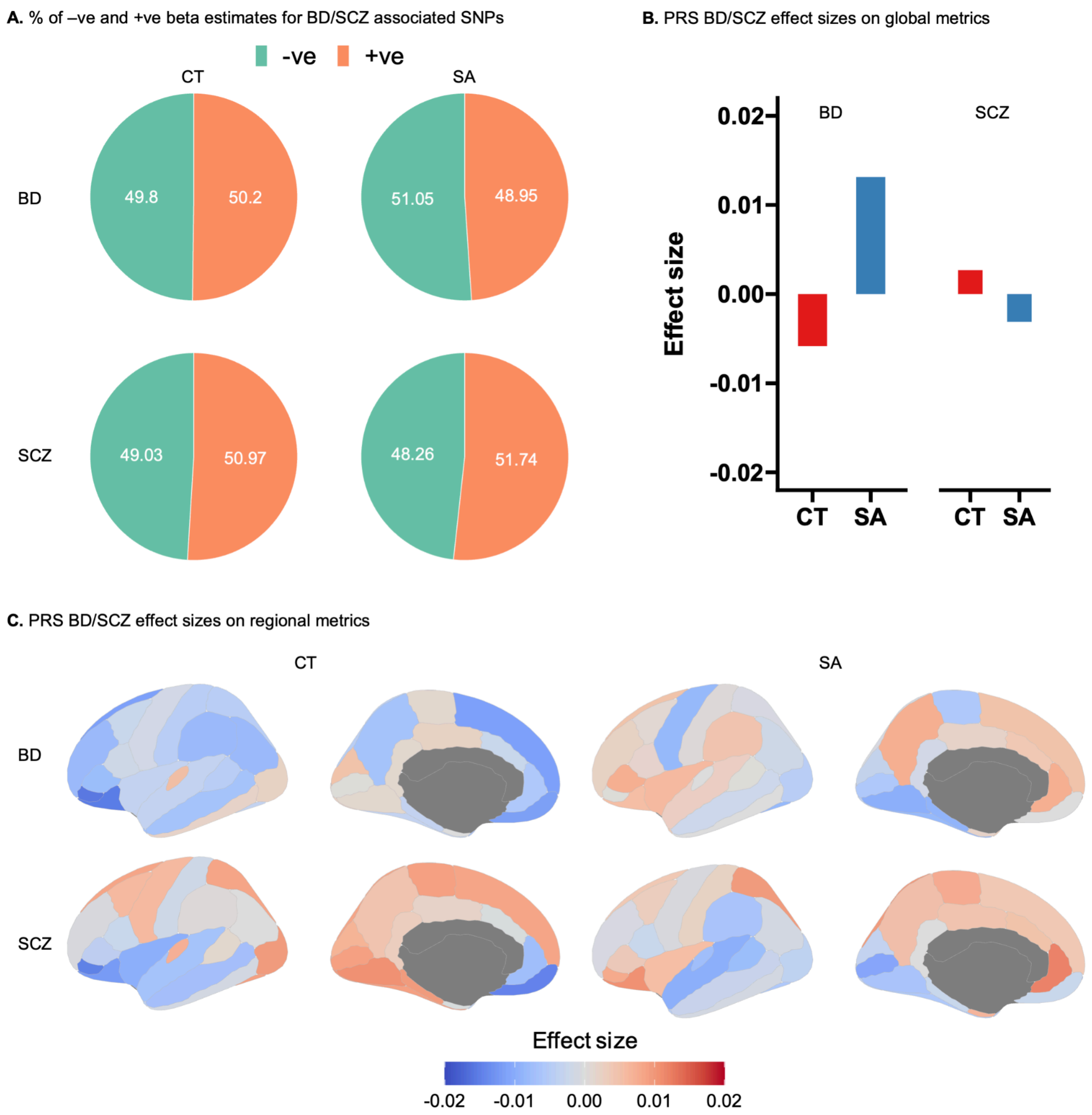


#### Supplement Figure 9. Cortical alterations across BD and SCZ associated common variants and polygenic risk scores (PRS).

Legend: A) Percentage of negative (-ve) and positive (+ve) regression estimates (derived from cortical structure GWAS for mean CT and total SA[13]) for BD/SCZ genome-wide associated SNPs. **B**-**C**) CT and SA effect sizes for BD and SCZ polygenic risk scores (PRS) for **B**) global, and **C**) regional measures. Notably, none of the PRS effect sizes were FDR significant (panels B and C). Effect sizes for PRS were derived using a linear regression in

31,000 UK Biobank participants of European ancestry, adjusting for age, sex, site, and ancestry principal components. Regional SA effect sizes were adjusted for total SA.

Abbreviations: BD=bipolar disorder; CT=cortical thickness; PRS=polygenic risk scores; SA: surface area; SCZ=schizophrenia.

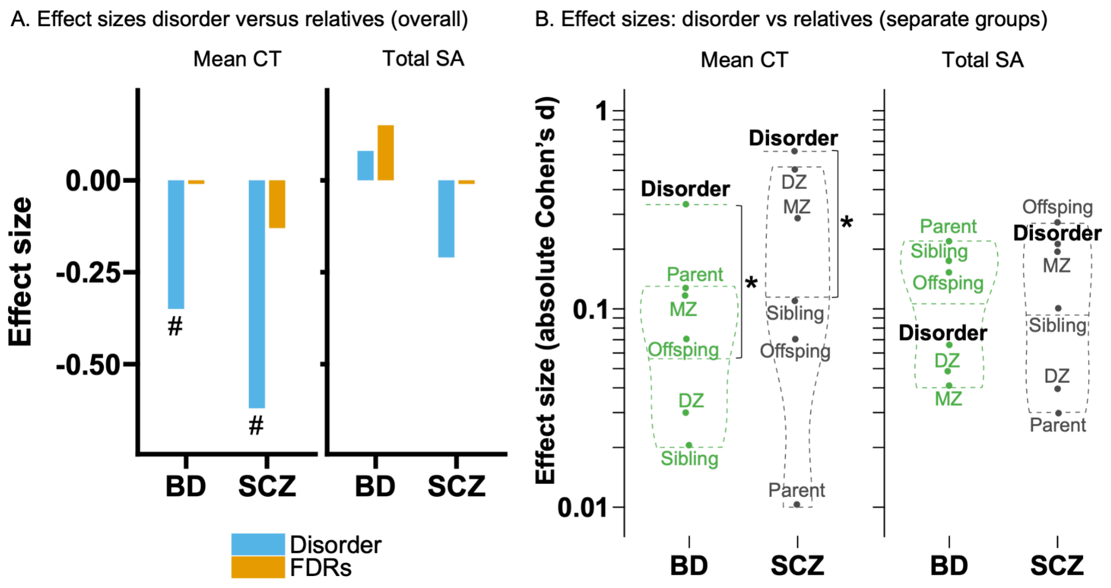


#### Supplement Figure 10. Diagnosis and first-degree relatives' comparison

Legend: **A**) Barplots comparing the effect sizes of disorder (BD or SCZ) with the first-degree relatives (FDRs). Data from ENIGMA relatives study[14] on BD and SCZ. #: FDR significant, as reported in the ENIGMA relatives study. **B**) Violin plots comparing the absolute effect sizes for mean CT and total SA between individuals with NPD diagnosis (disorder) and their first-degree relatives, separated by relative categories. *: FDR q-value <0.05, using non-parametric one-sample Wilcoxon signed-rank test, assessing if the median of the first-degree relatives' effect sizes were less than disorder effect size.

Abbreviations, BD=bipolar disorder; DZ=di-zygotic twin; MZ=mono-zygotic twin; SCZ=schizophrenia.
